# A Systematic Review on the Effects of Exercise on Gut Microbial Diversity, Taxonomic Composition, and Microbial Metabolites: Identifying Research Gaps and Future Directions

**DOI:** 10.1101/2023.09.05.23294991

**Authors:** John M.A. Cullen, Shahim Shahzad, Jaapna Dhillon

## Abstract

The gut microbiome, hosting a diverse microbial community, plays a pivotal role in metabolism, immunity, and digestion. While the potential of exercise to influence this microbiome has been increasingly recognized, findings remain incongruous. This systematic review examined the effects of exercise on the gut microbiome of human and animal models. Databases (i.e., PubMed, Cochrane Library, Scopus, and Web of Science) were searched up to June 2022. Thirty-two exercise studies, i.e., 19 human studies, and 13 animal studies with a minimum of two groups that discussed microbiome outcomes, such as diversity, taxonomic composition, or microbial metabolites, over the intervention period, were included in the systematic review (PROSPERO registration numbers: CRD42023394223 and CRD42023394485). Results indicated that over 50% of studies found no significant exercise effect on human microbial diversity. When evident, exercise often augmented the Shannon index, reflecting enhanced microbial richness and evenness, irrespective of disease status. Changes in beta-diversity metrics were also documented with exercise but without clear directionality. A larger percentage of animal studies demonstrated shifts in diversity compared to human studies, but without any distinct patterns, mainly due to the varied effects of predominantly aerobic exercise on diversity metrics. In terms of taxonomic composition, in humans, exercise usually led to a decrease in the Firmicutes/Bacteroidetes ratio, and consistent increases with *Bacteroides* and *Roseburia* genera. In animal models, *Coprococcus*, a SCFA producer, consistently rose with exercise. Generally, SCFA producers were found to increase with exercise in animal models. With regard to metabolites, SCFAs emerged as the most frequently measured metabolite. However, due to limited human and animal studies examining exercise effects on microbial-produced metabolites, including SCFAs, clear patterns did not emerge. The overall risk of bias was deemed neutral. In conclusion, this comprehensive systematic review underscores that exercise can potentially impact the gut microbiome with indications of changes in taxonomic composition. The significant variability in study designs and intervention protocols demands more standardized methodologies and robust statistical models. A nuanced understanding of the exercise-microbiome relationship could guide individualized exercise programs to optimize health.

## 1. Introduction

The gut microbiome is a complex and diverse ecosystem of microorganisms that plays a vital role in various physiological processes, including digestion, immunity, and metabolism. In recent years, the potential of exercise to modulate the gut microbiome has gained considerable attention, and numerous studies have investigated the effects of exercise on the gut microbiome in both human and animal models.

The first indications that exercise has a role in modulating the human gut microbiome and metabolic capacity of the microbiota were observed through cross-sectional studies (Mailing et al., 2019). For example, professional rugby players were found to have greater alpha diversity (Clarke et al., 2014), and amino acid, carbohydrate metabolic pathways, and greater fecal SCFAs (Barton et al., 2018) than sedentary individuals. Moreover, even in the non-athletic populations, women who exercised for at least three hours per wk had increased levels of butyrate producers such as *Faecalibacterium prausnitzii, Roseburia hominis*, and mucin degrader, *Akkermansia muciniphila*. However, the cross-sectional nature of these studies limits the ability of these studies to control diet and other confounding factors on the microbiome (Mailing et al., 2019).

The mechanisms by which exercise can impact the gut microbiome are not yet fully understood, but several potential mechanisms have been proposed (Mailing et al., 2019). The gut-associated lymphoid tissue (GALT) houses a majority of our immune cells, and exercise can modulate their gene expression, favoring anti-inflammatory and antioxidant profiles (Hoffman-Goetz et al., 2009, 2010; Packer and Hoffman-Goetz, 2012), potentially mediating microbial host-microbial interactions (Ismail et al., 2011; Mailing et al., 2019). Exercise can also influence the gut’s mucus layer, a critical barrier between microbes and the gut lining; as well as gut motility which can alter GI transit time, potentially impacting microbial habitats and their nutrient availability (Mailing et al., 2019). Furthermore, exercise can modify the circulation of bile acids (Meissner et al., 2011), known regulators of the microbial community structure (Kakiyama et al., 2013; Mailing et al., 2019). Finally, the metabolic demands of exercise release various compounds like lactate and myokines (Egan and Zierath, 2013) that might directly or indirectly interact with the gut environment (Mailing et al., 2019) potentially influencing the microbiome. Overall, the impact of exercise on the gut microbiome is likely mediated by a combination of these mechanisms. However, the magnitude and consistency of these effects are still unclear, and there is significant variation in study designs, including differences in exercise types, durations, intensities, and populations studied, as well as confounding effects of uncontrolled dietary and environmental factors.

To address these issues, this systematic review aims to synthesize the available data on the effects of exercise on gut microbiome in humans and animal models. The study of the impact of exercise on the animal microbiome is important because it can provide valuable mechanistic insights into the complex interactions between host physiology, behavior, and the gut microbiota. Additionally, animal models allow for greater experimental control of variables such as diet and genetics, which can be difficult to control in human studies. More specifically, the systematic review examines the impact of exercise type, duration, and intensity on changes in gut microbiome outcomes such as microbial phyla and genera composition, alpha- and beta-diversity, and microbial-produced metabolites such as SCFAs etc., in trials with 2 or more arms, in both human and animal models. The results of this review will provide a better understanding of the relationship between exercise and the gut microbiome, potentially identifying targets for interventions to improve gut and metabolic health.

## 2. Methods

The study protocols for the human and animal reviews are registered with PROSPERO, the International Prospective Register of Systematic Reviews (CRD42023394223 and CRD42023394485)

### Search Strategy

A search strategy was designed based on guidelines in the Cochrane Handbook of systematic reviews (Cochrane Handbook for Systematic Reviews of Interventions, n.d.). Specific terms and language were utilized to classify and analyze studies that included exercise and assessment of the gut microbiome. Search strategies are listed in **Supplemental Table 1.** Primary searches were conducted until October 29, 2021, and were then updated from October 2021 to June 1, 2022 to obtain studies that were published in the interim. There are no restrictions on language, study design, or date of publication.

### Study Selection

The study search was performed across 4 databases, PubMed, Web of Science, Scopus, and Cochrane. Results were then collected in Zotero (version 5.0.87). To determine inclusion of studies in the review, three authors, (JD, JC, SS) reviewed the articles in a methodical manner. Any disparities were decided through an inclusion vote.

In the first pass, titles were independently screened to identify relevant articles based on specific criteria. Articles were excluded if the studies were 1) duplicates, 2) reviews, perspectives, or protocol papers, 3) published as conference abstracts, 4) not related to either exercise or the gut microbiome, or 5) were in children or pregnant populations. In the second pass, abstracts were screened for the same criteria in the first pass with additional criteria to exclude studies that were 1) either observational or cross-sectional, 2) not exercise interventions, and 3) did not have full texts. In the third pass, full texts were screened for the same criteria in the first two passes with additional criteria to exclude studies that 1) did not include comparator groups, 2) collected gut microbiome data at one time point, 3) studies where exercise interventions were not clearly defined. Lastly, articles were categorized based on human studies and animal studies (mouse, rat, and horse).

### Data Extraction

Data collection tables were developed and finalized by 3 authors (JD, JC, SS). Extraction of data was performed by 2 authors (JC, SS) and was reviewed for accuracy by JD. Study quality was evaluated using the Oxford Centre for Evidence-Based Medicine Levels of Evidence (Oxford Centre for Evidence-Based Medicine: Levels of Evidence (March 2009) — Centre for Evidence-Based Medicine (CEBM), University of Oxford, n.d.). The risk of bias analysis was conducted using the quality control checklists of the Academy of Nutrition and Dietetics (Academy of Nutrition and Dietetics, n.d.).

## 3. Results

In total, 4731 records were screened for inclusion from database searches and 35 from meta-analyses and systematic reviews and manual search of reference lists. A total of 4608 records were excluded based on the established criteria (**Figure 1**). Thirty-two studies, i.e., 19 human studies, and 13 animal studies with 2 or more groups that discussed microbiome outcomes, such as diversity, taxonomic composition, or microbial metabolites, over the intervention period, were included in the systematic review. The detailed inclusion and exclusion criteria are presented in **Figure 1**. The breakdown of outcomes by exercise intervention type is depicted in **Figure 2**.

**Figure 1:**
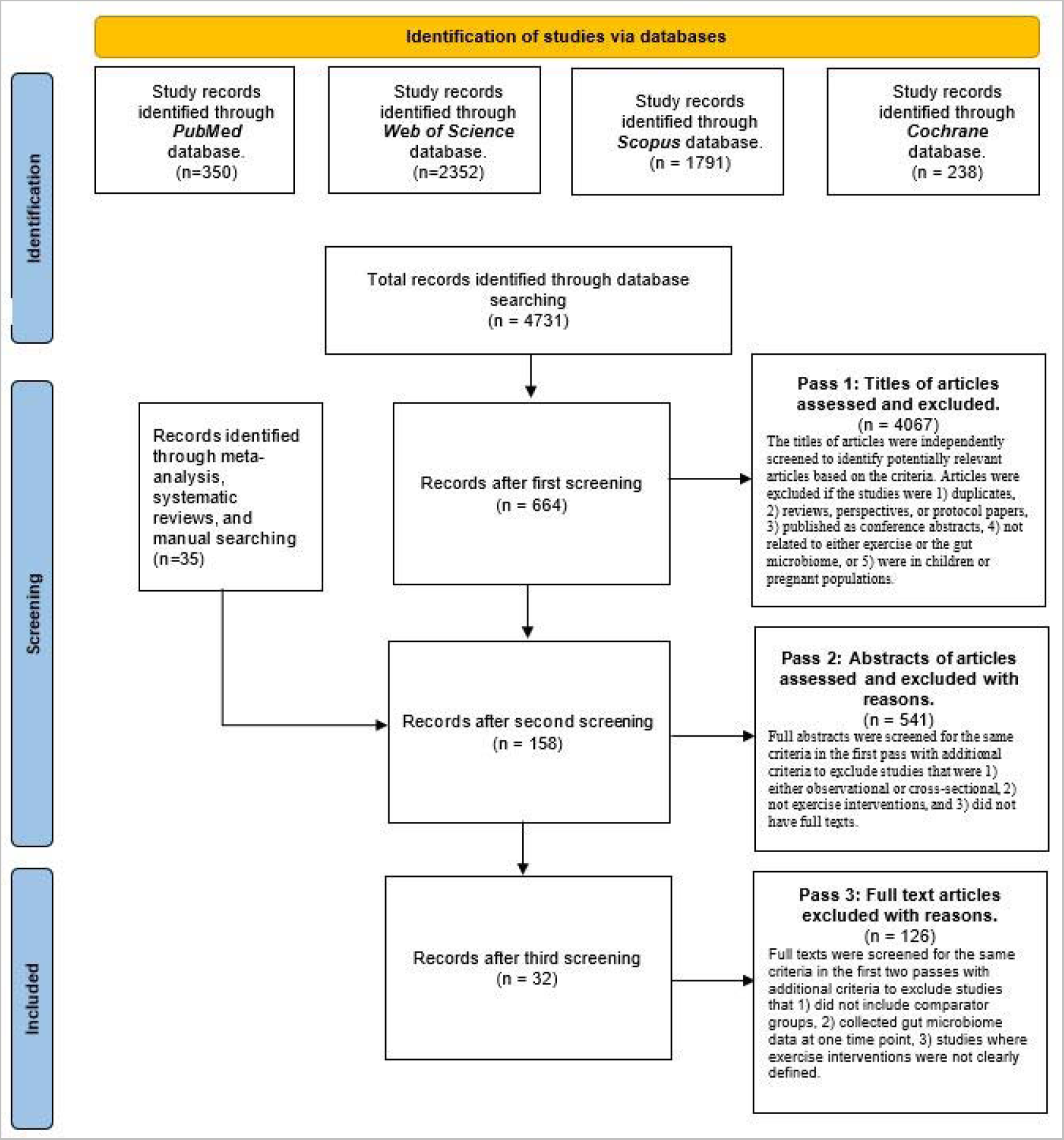
PRISMA flow diagram for inclusion of studies in the systematic review. PRISMA, Preferred Reporting Items for Systematic Reviews and Meta-Analyses.

**Figure 2:**
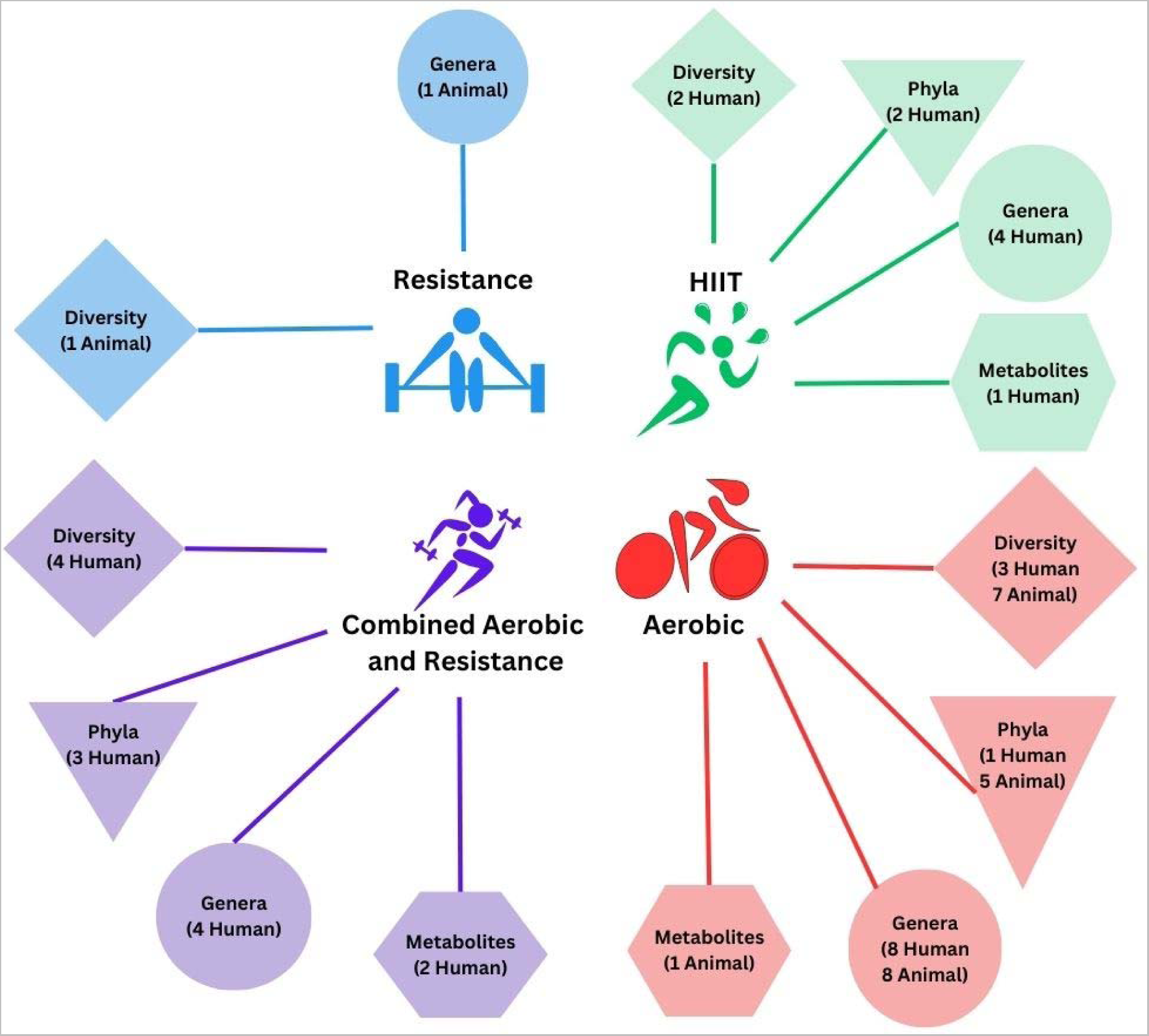
Microbiome outcomes stratified by exercise intervention type. The number indicates the number of studies that demonstrate a change in response to the exercise intervention in humans or animal models. Direction of change is not quantified. Studies incorporating HIIT are stratified separately regardless of whether the study included other exercise components in addition to HIIT.

### Intervention type, study design, duration, and population profiles

Nineteen studies conducted in humans met our inclusion criteria. Of those, 11 included aerobic training interventions (Taniguchi et al., 2018; Morita et al., 2019; Resende et al., 2021; Moitinho-Silva et al., 2021; Mahdieh et al., n.d.; Cheng et al., 2012; Sun et al., 2019; Calabrese et al., 2022; Motiani et al., 2020; Kern et al., 2020; Warbeck et al., 2021), 2 included resistance training interventions (Morita et al., 2019; Moitinho-Silva et al., 2021) and 9 included combined aerobic and resistance training interventions (Cronin et al., 2018a, 2019; Liu et al., 2020; Mokhtarzade et al., 2021; Zhong et al., 2021; Calabrese et al., 2022; Dupuit et al., 2022; Torquati et al., 2022; Wei et al., 2022). Six studies included a high intensity interval training intervention (Liu et al., 2020; Motiani et al., 2020; Warbeck et al., 2021; Dupuit et al., 2022; Sun et al., 2022; Torquati et al., 2022) Included studies varied in design, duration, and populations studied (**Table 1**). Seventeen studies were trials with a parallel design (2–6 arms) (Cronin et al., 2018a; Morita et al., 2019; Kern et al., 2020; Liu et al., 2020; Motiani et al., 2020; Moitinho-Silva et al., 2021; Mokhtarzade et al., 2021; Resende et al., 2021; Warbeck et al., 2021; Zhong et al., 2021; Calabrese et al., 2022; Cheng et al., 2022; Dupuit et al., 2022; Sun et al., 2022; Torquati et al., 2022; Wei et al., 2022; Mahdieh et al., n.d.), of which the longest trial had a duration of 12 mon (Wei et al., 2022) and the shortest trials had a duration of 2 wk (Motiani et al., 2020). One study followed a crossover design in which participants were exposed to a 5 wk exercise intervention and a 5 wk control (Taniguchi et al., 2018). Another study was an 8-wk RCT with a 2-arm parallel design with a partial crossover (Cronin et al., 2019). The studies involve a diverse range of participant profiles, including individuals with prediabetes or type 2 diabetes, older males and females, individuals with normal blood pressure and non-diabetic status, those with inflammatory bowel disease (IBD), multiple sclerosis (MS), nonalcoholic fatty liver disease (NAFLD), and celiac disease, as well as individuals with overweight and obesity (**Table 1**).

**Table 1.**
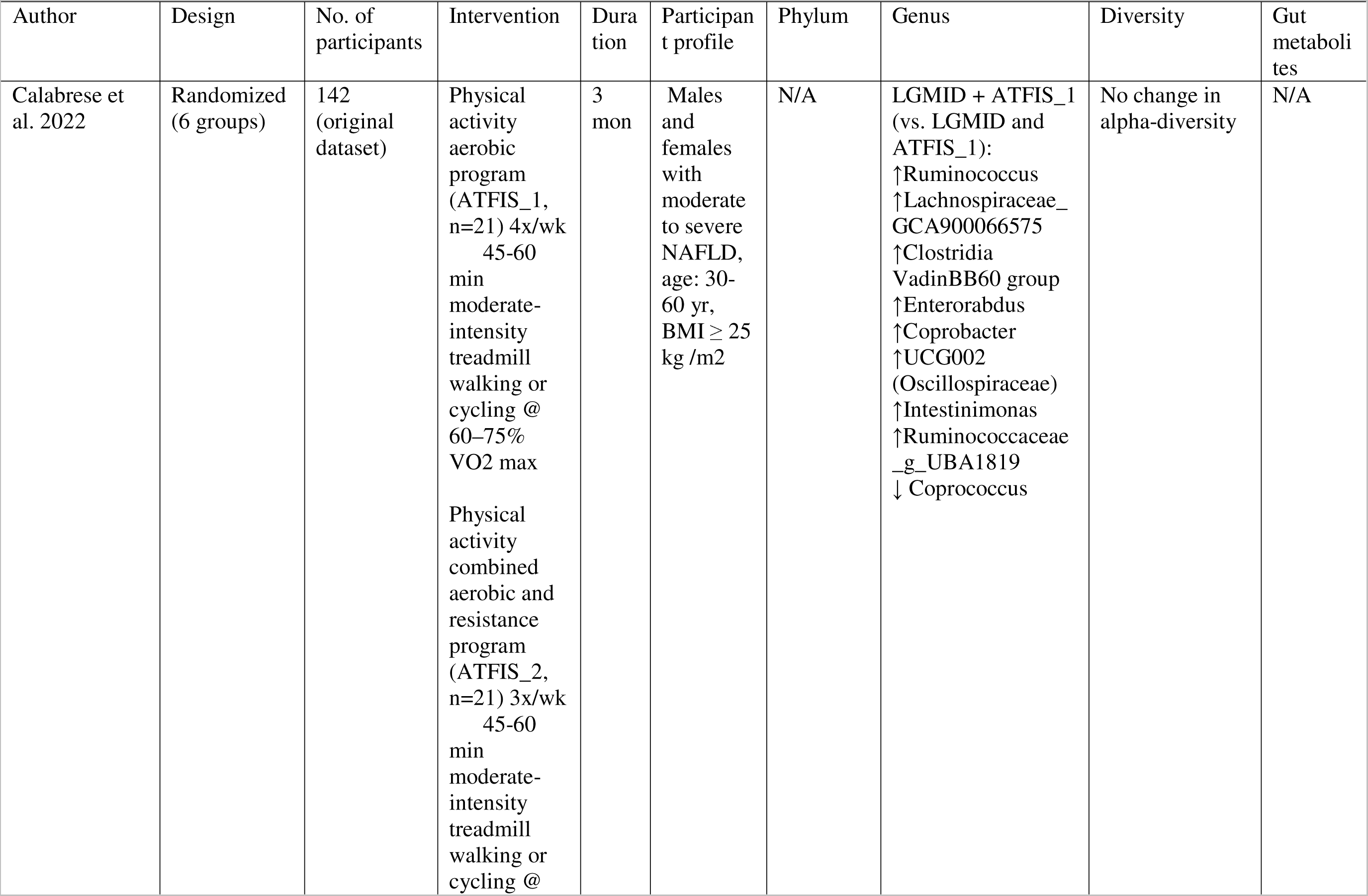
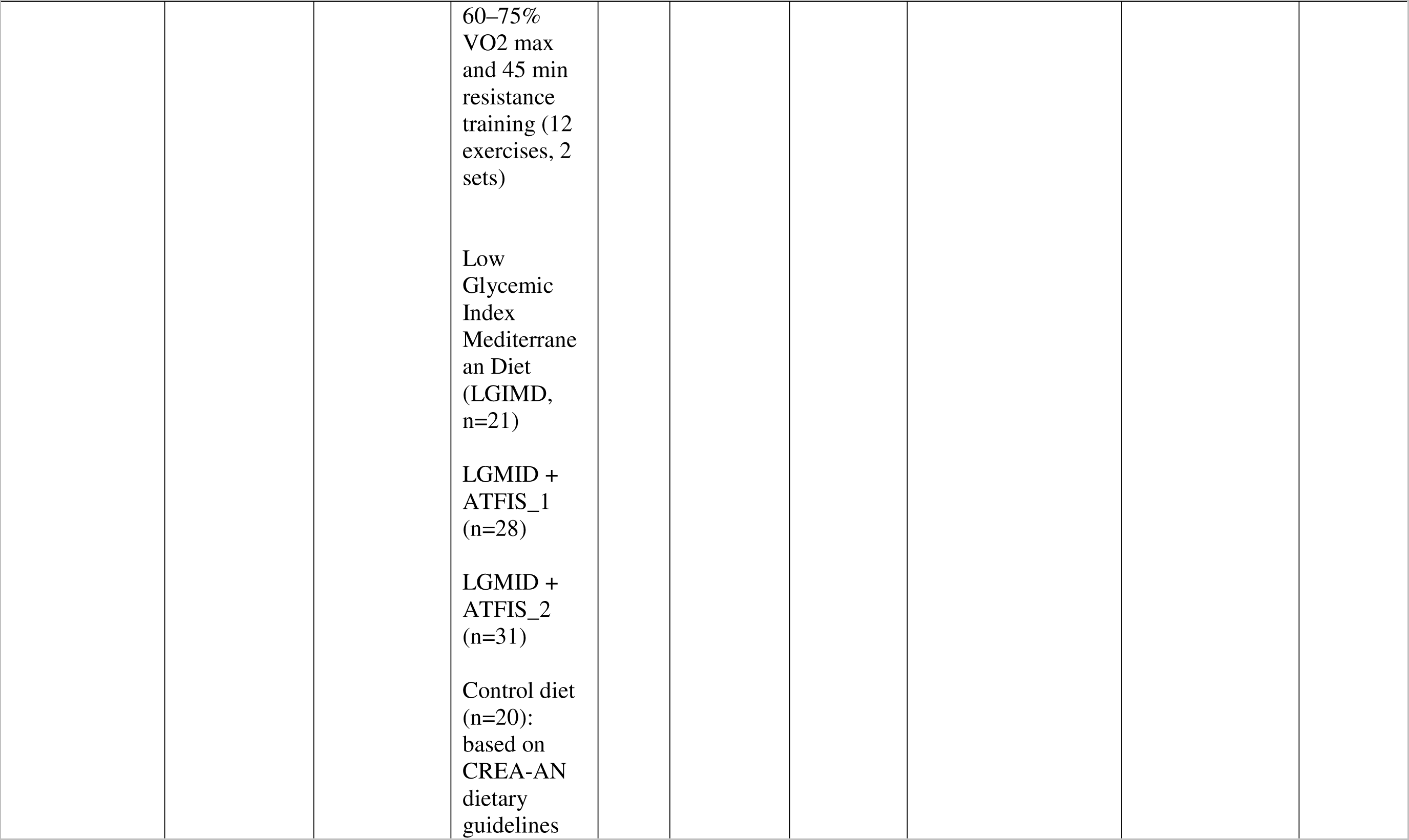
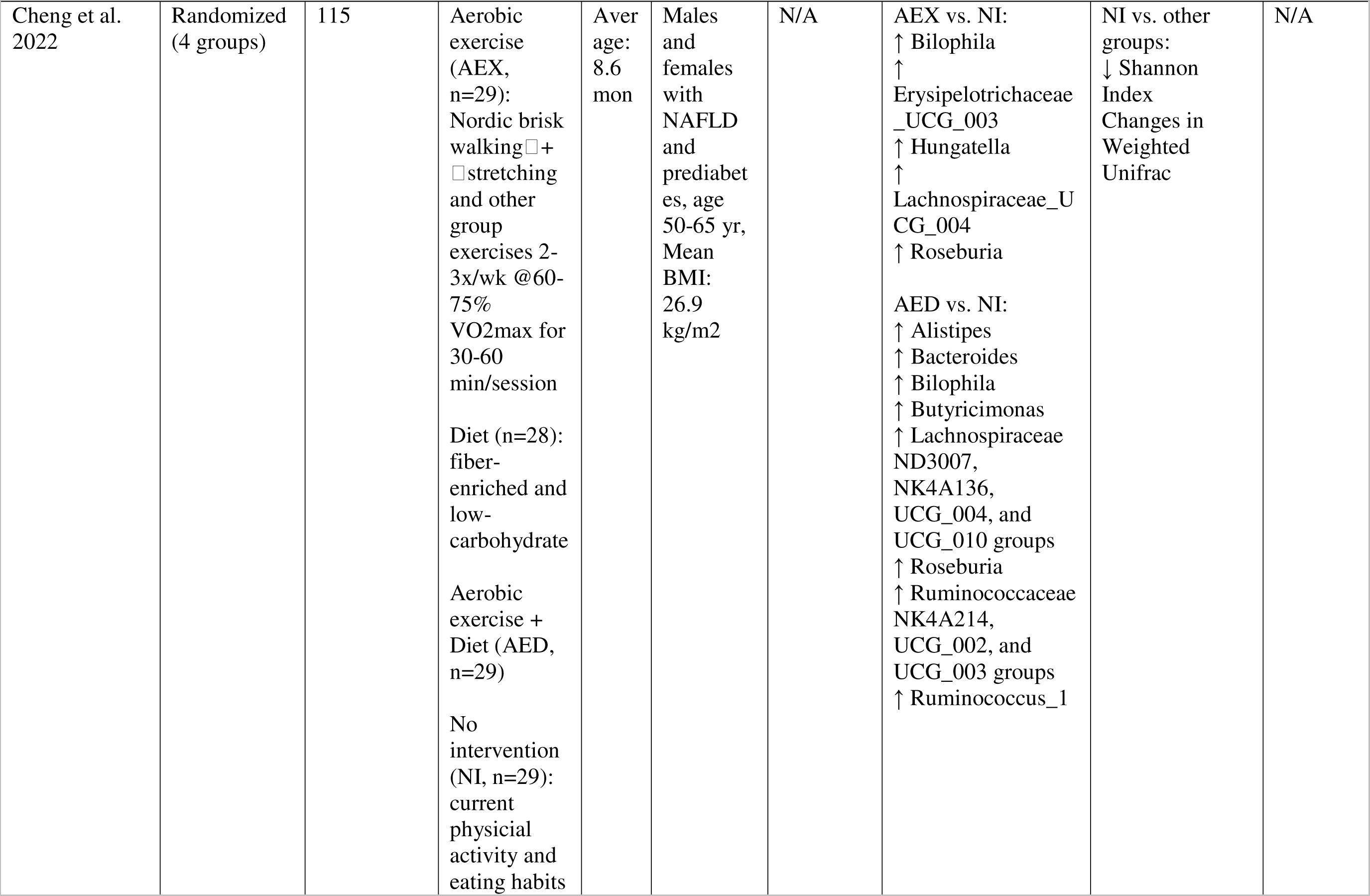
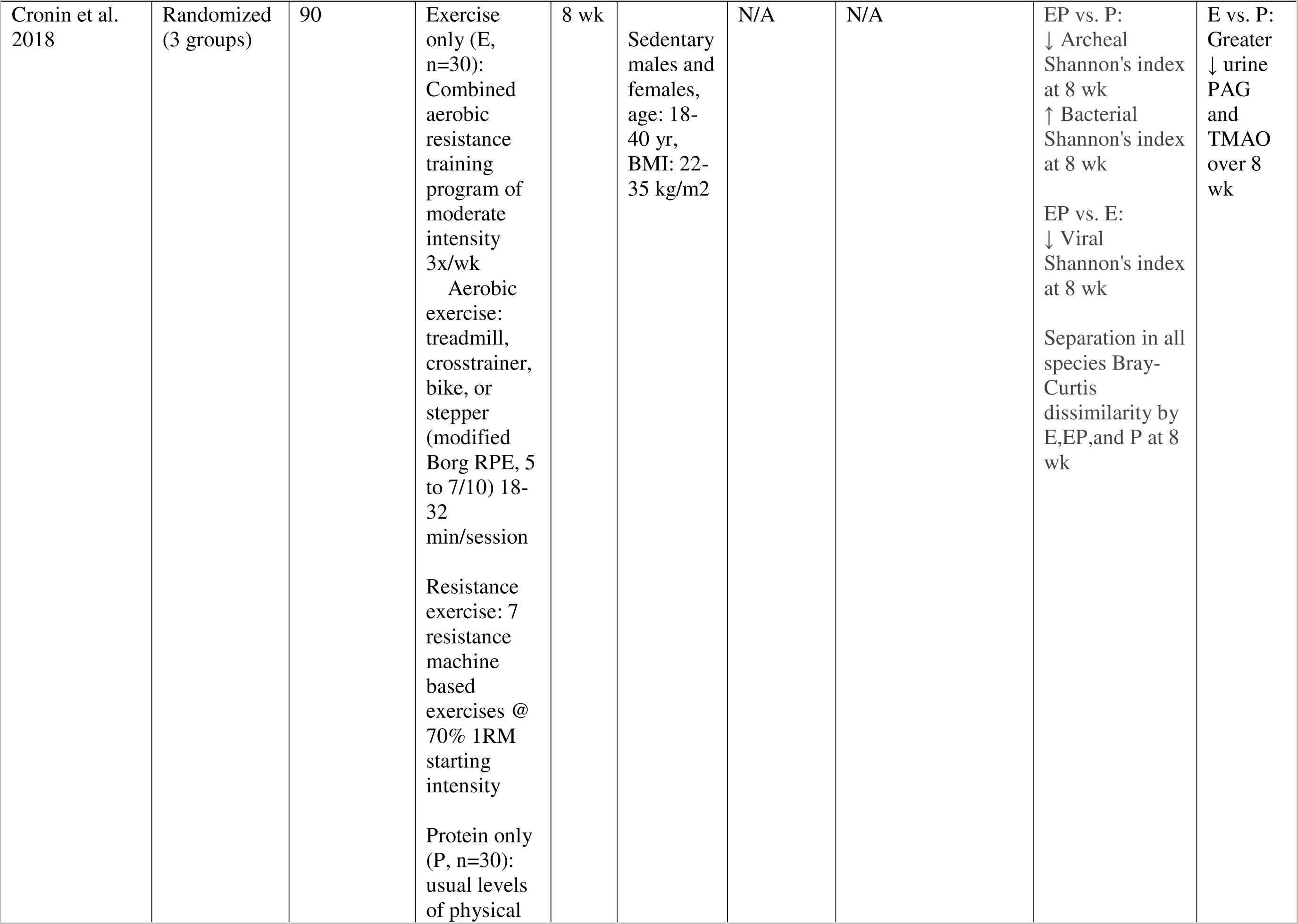
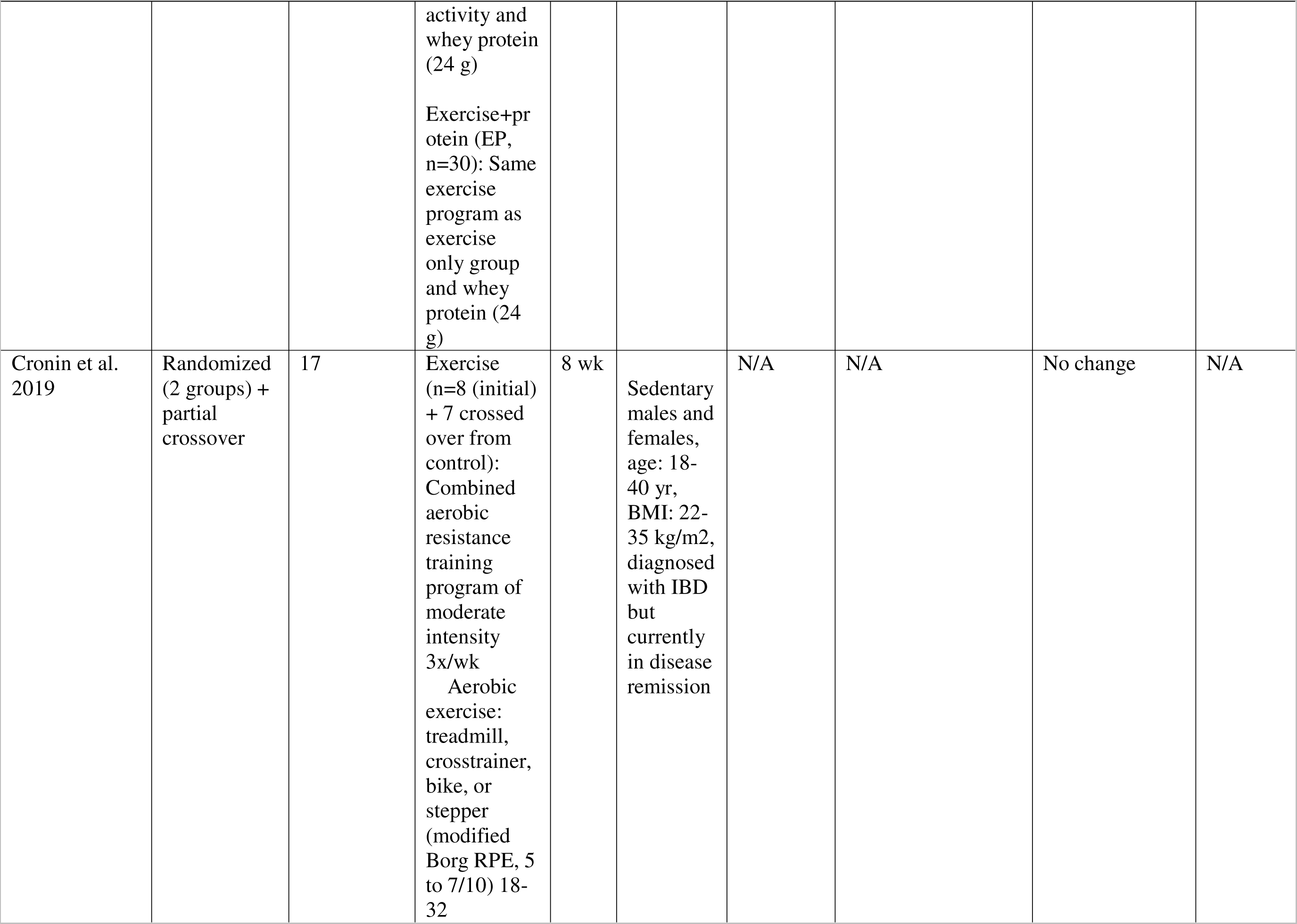
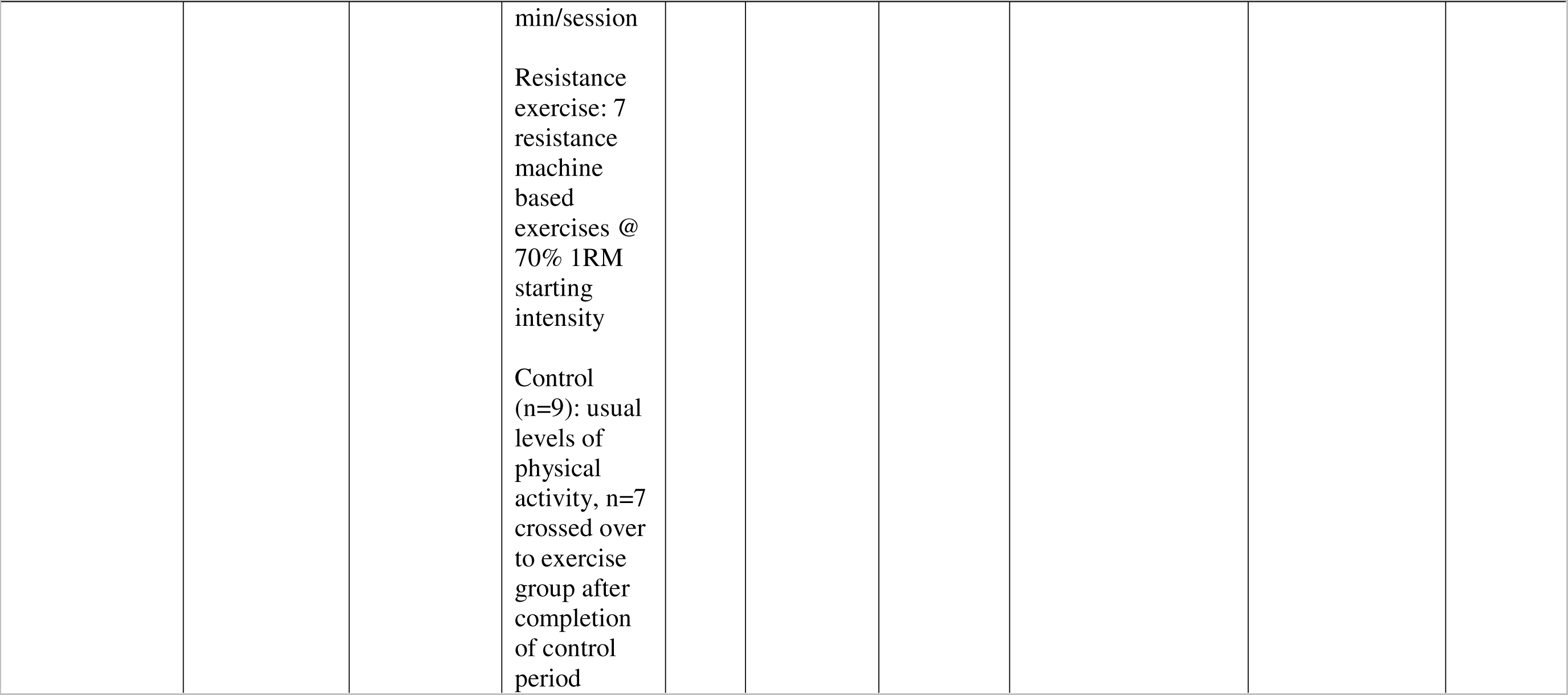
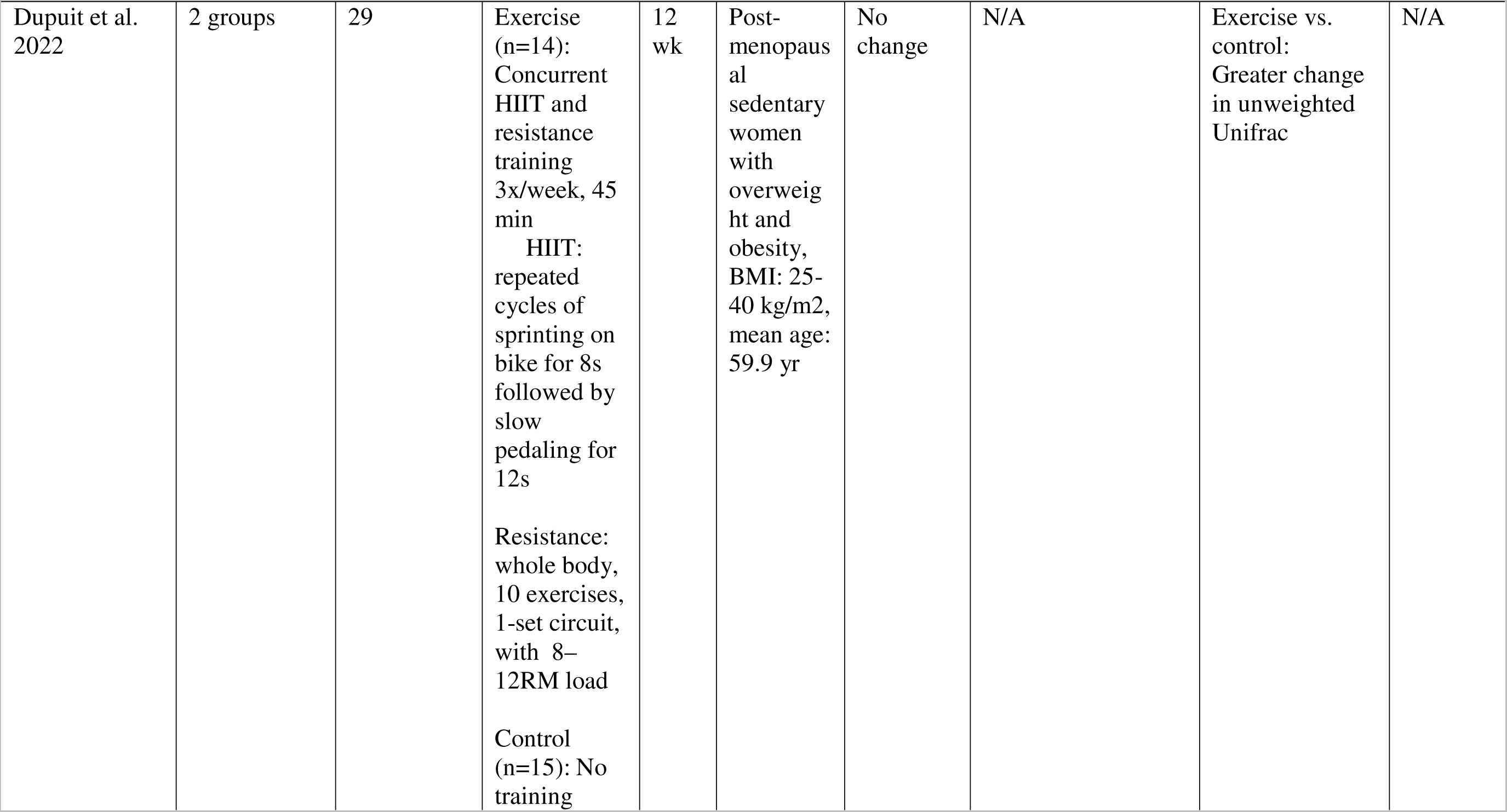
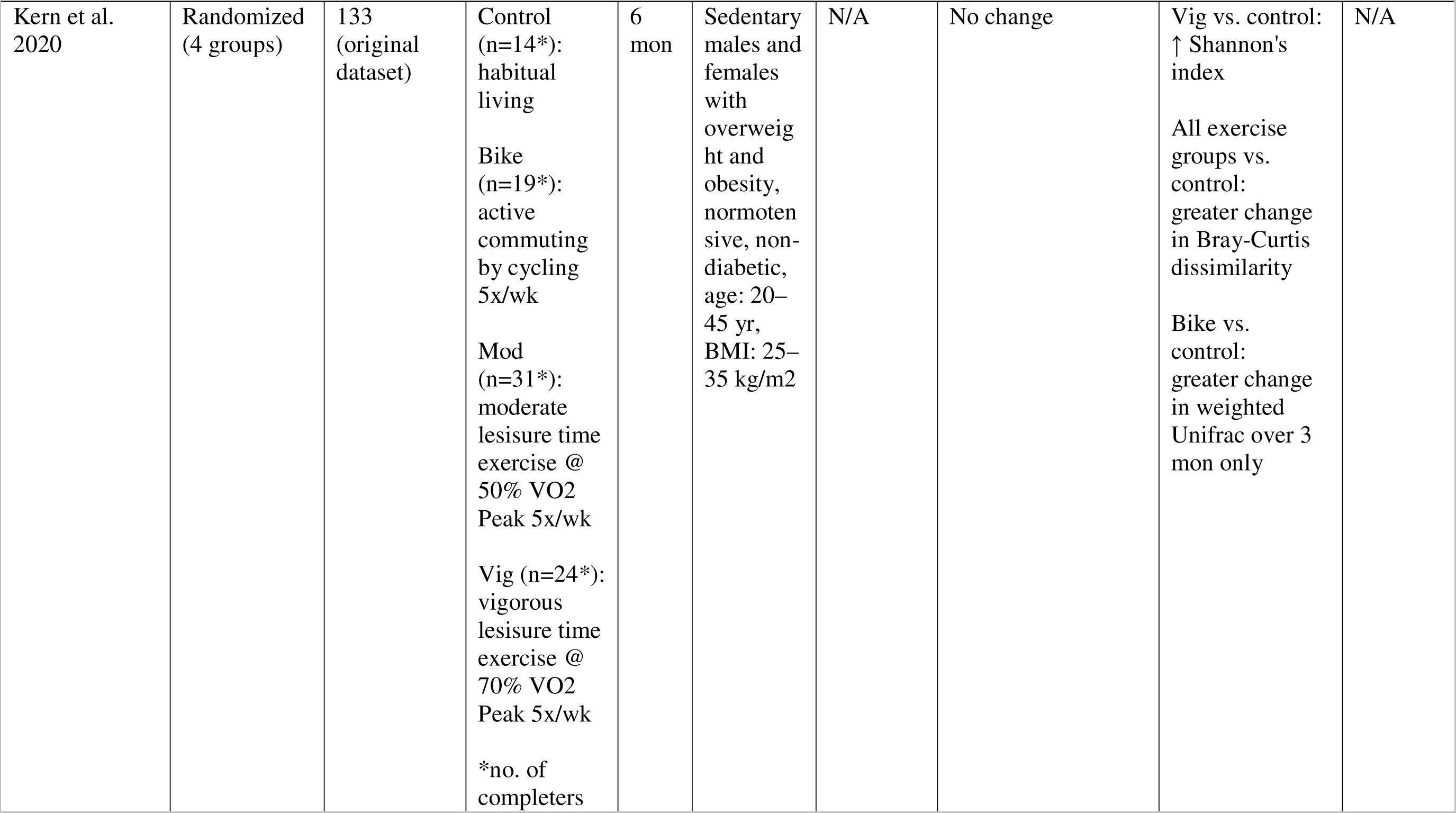
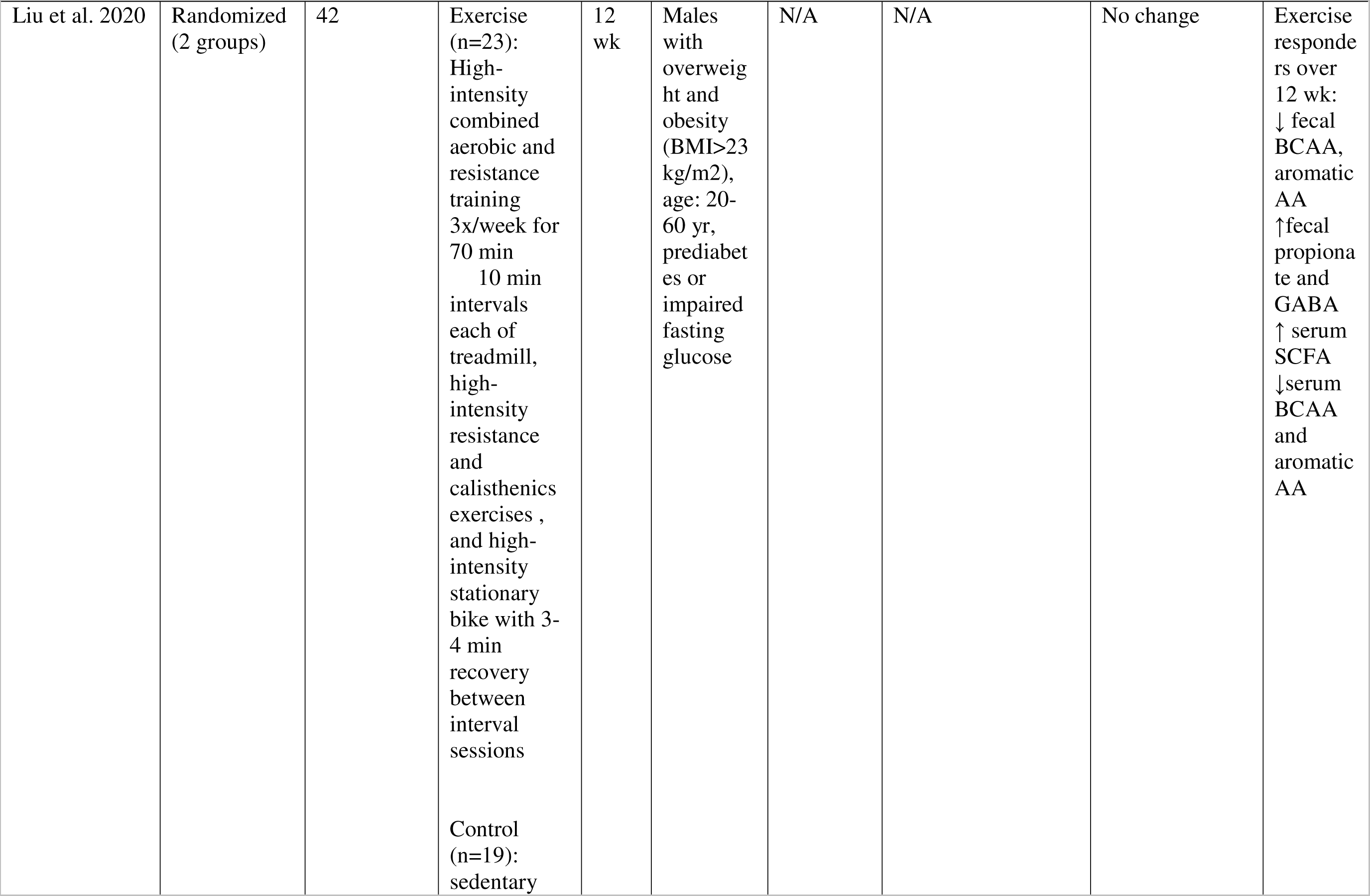
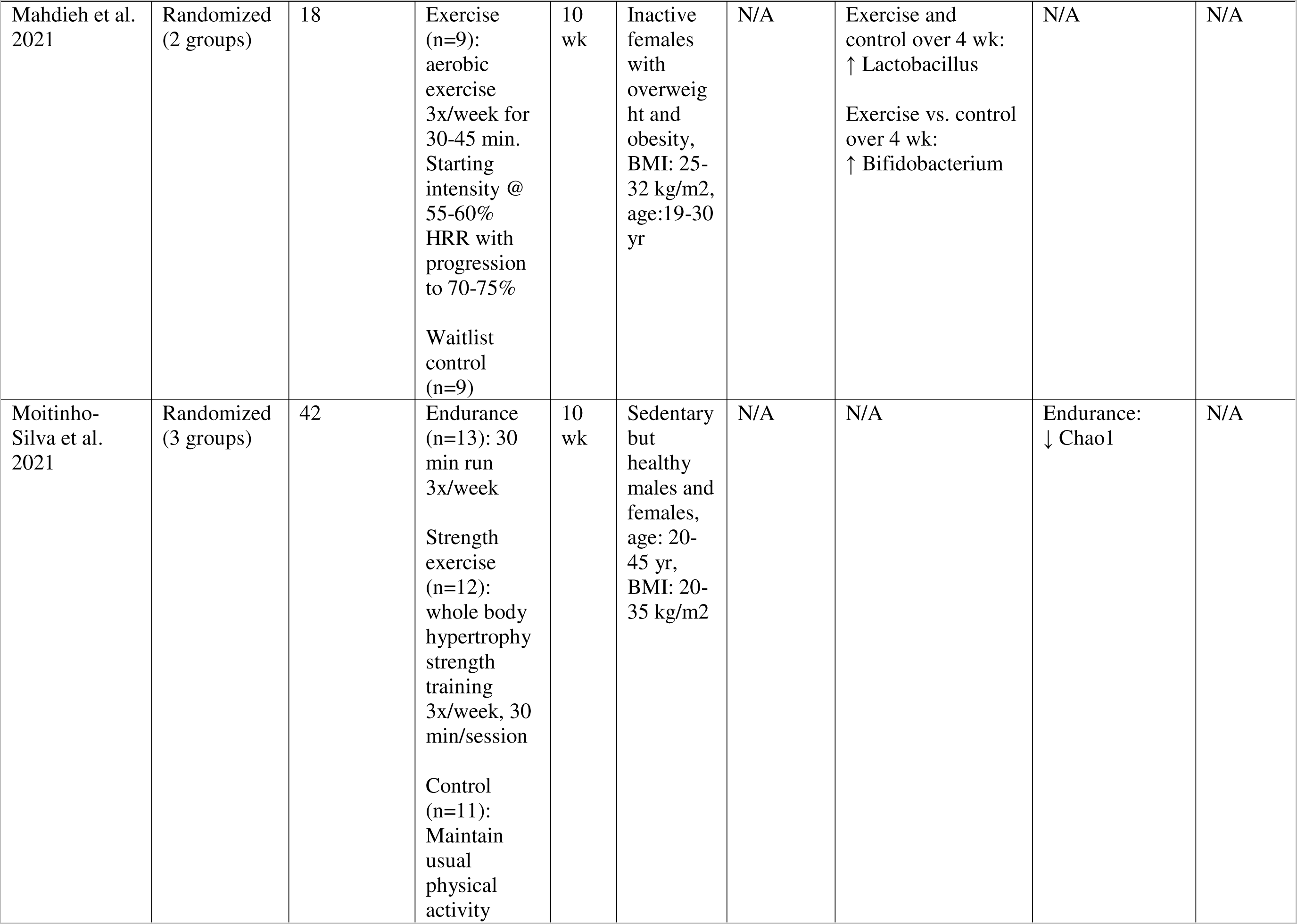
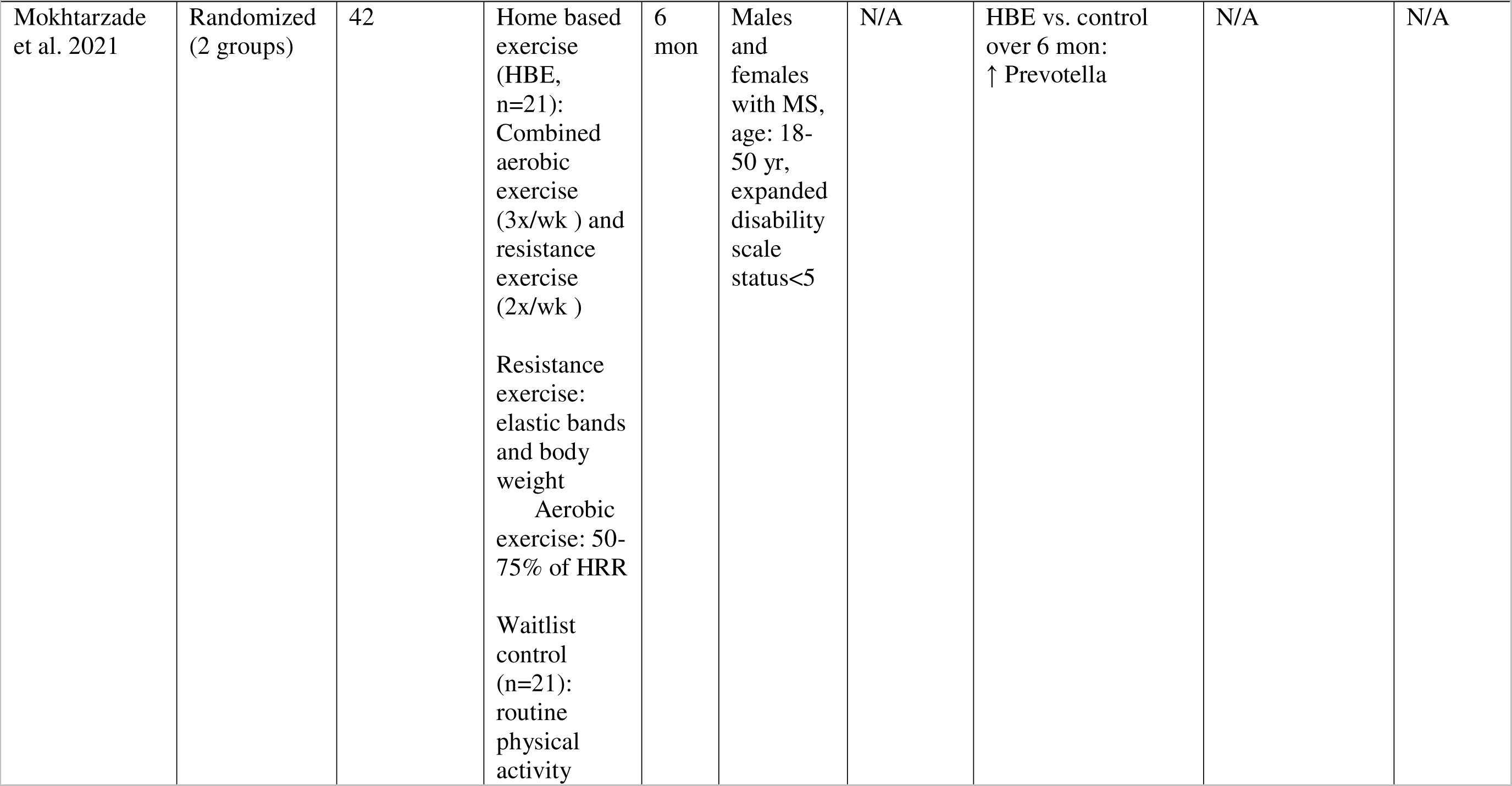
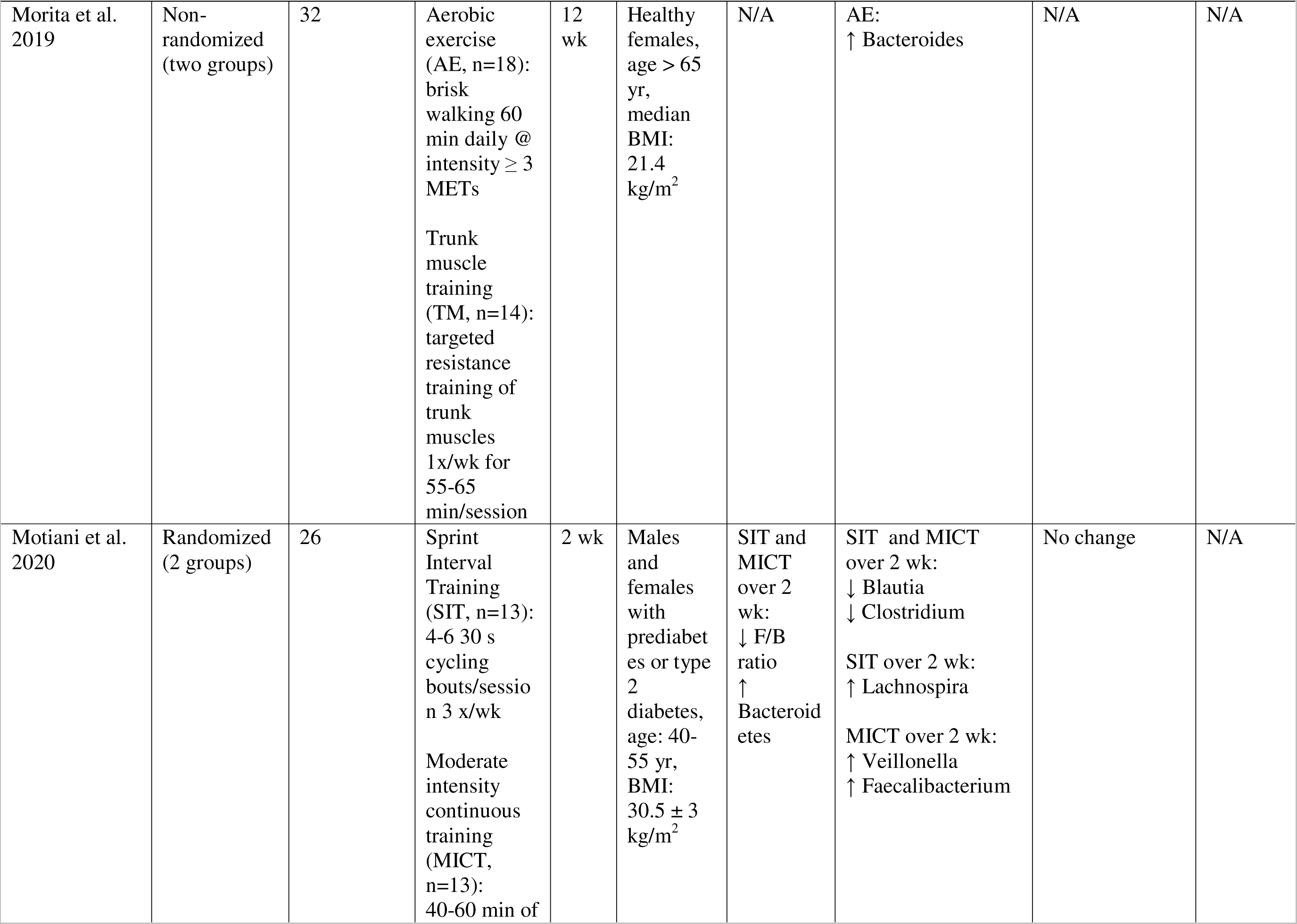
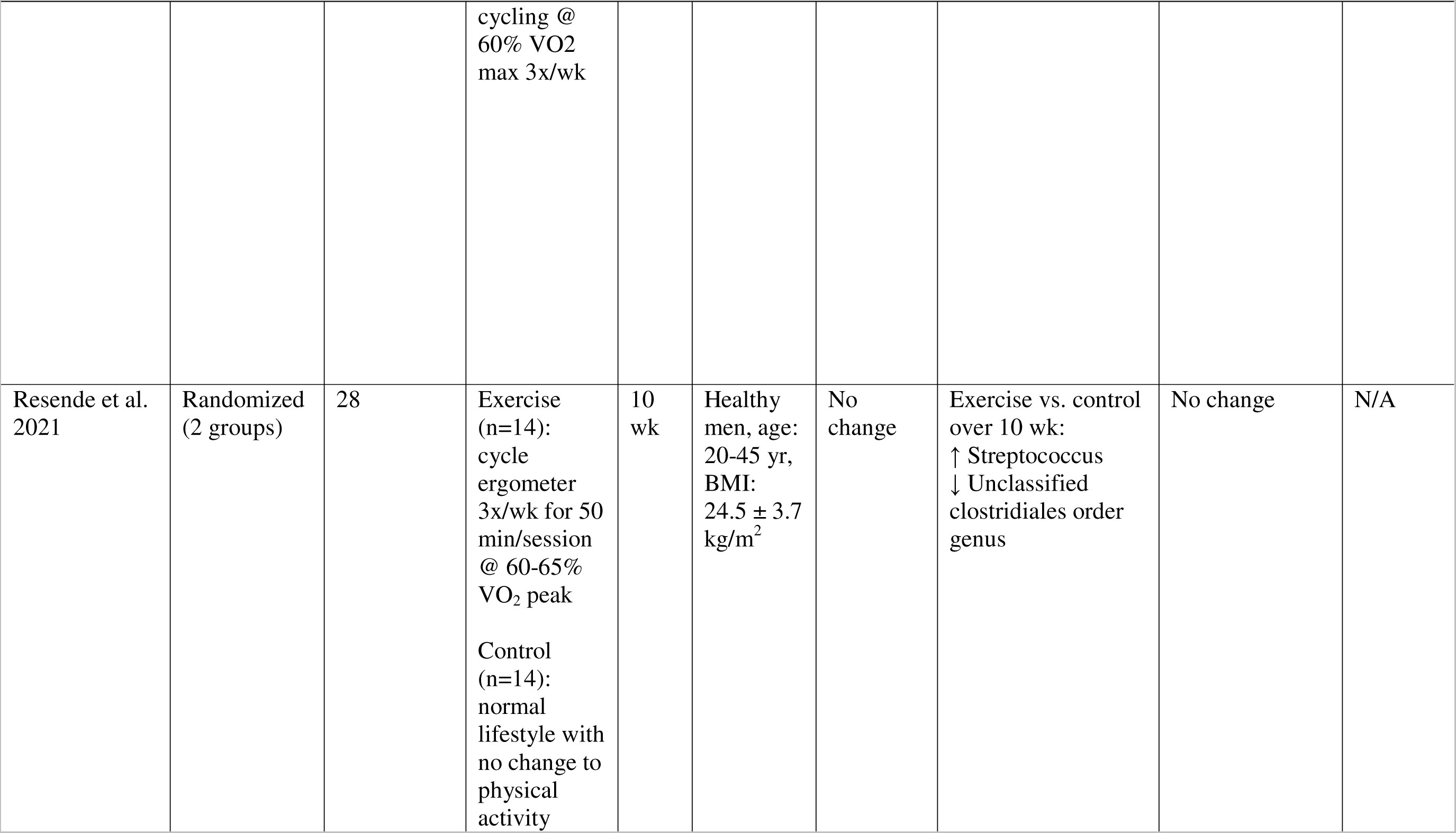
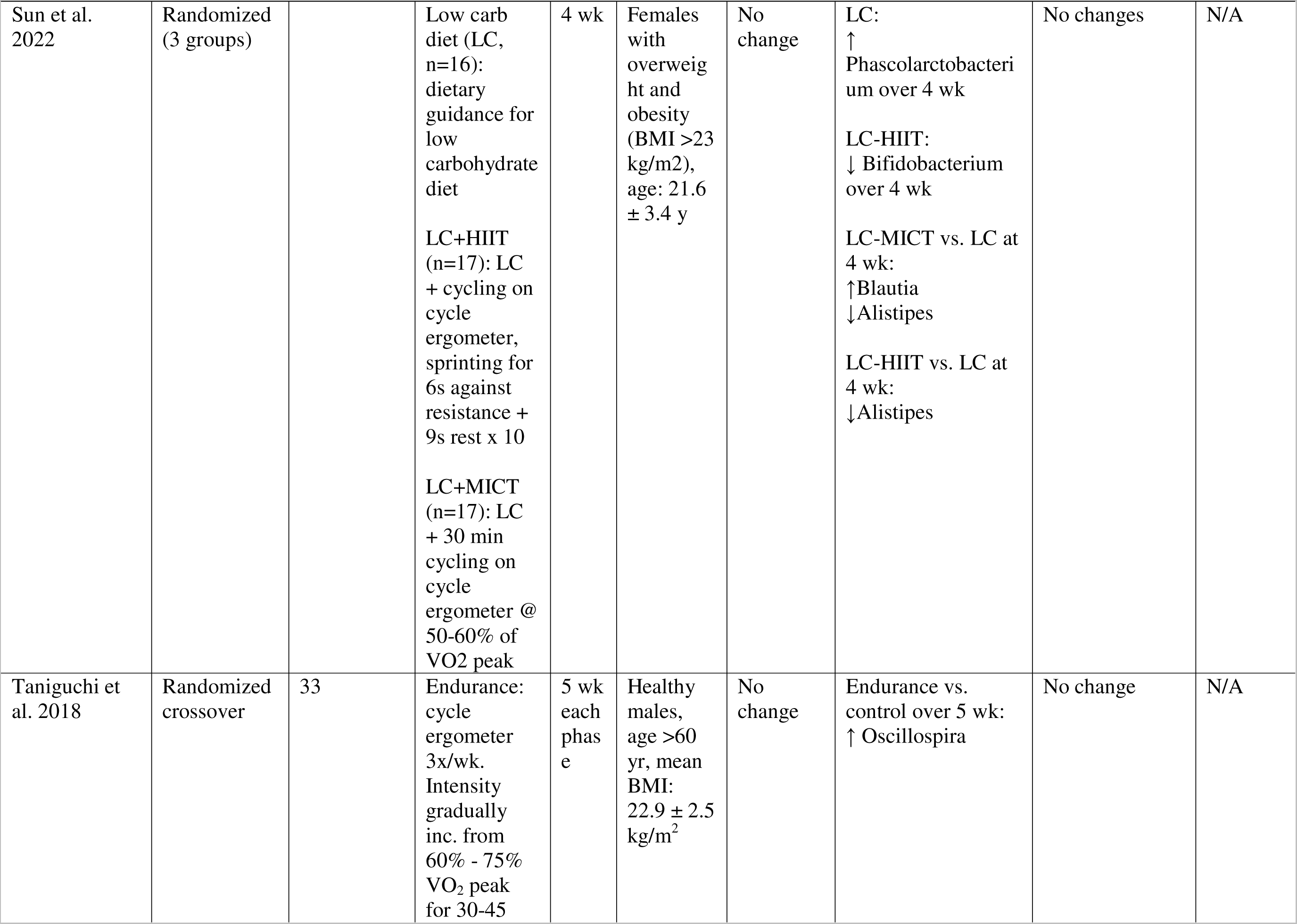
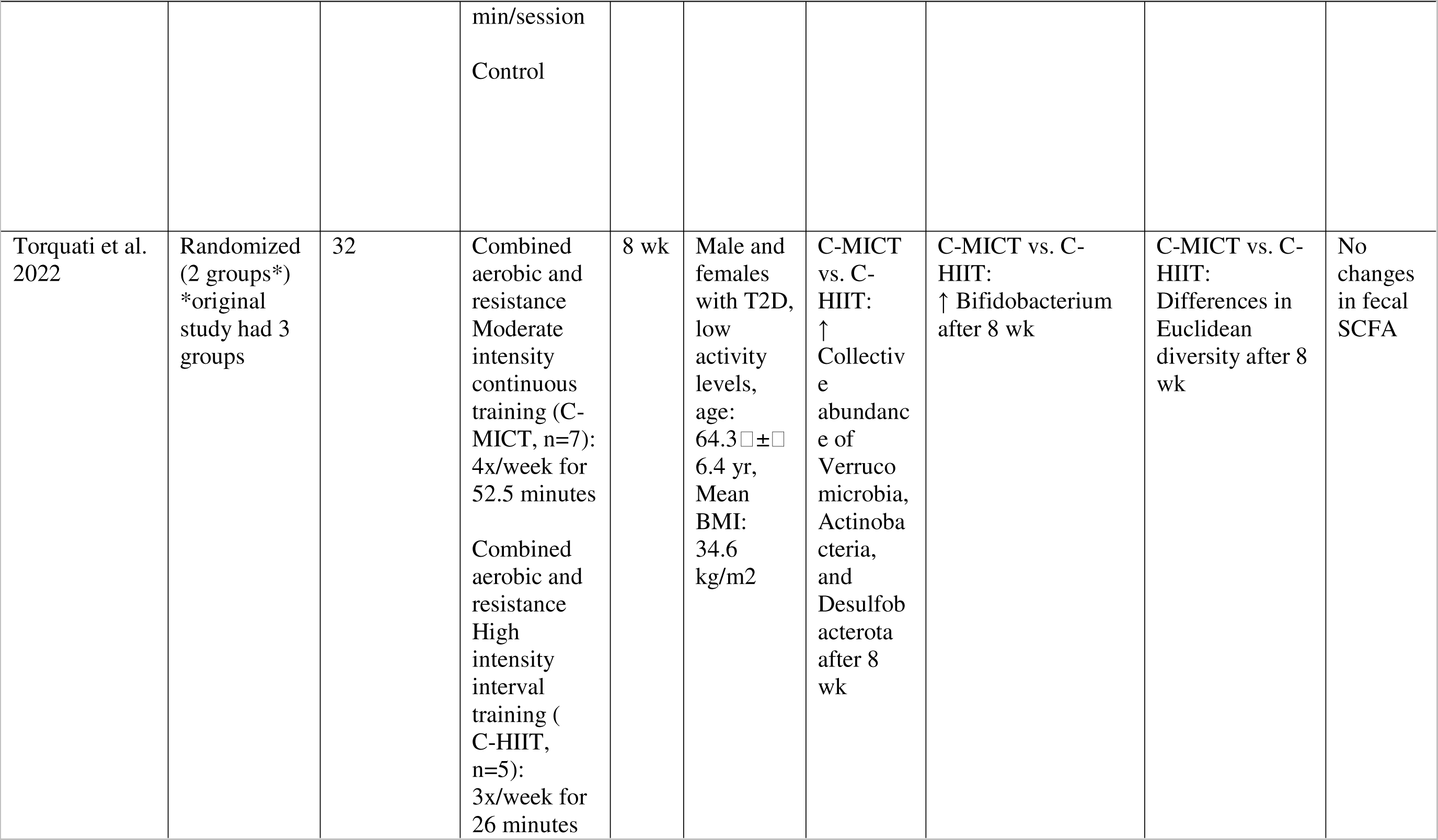
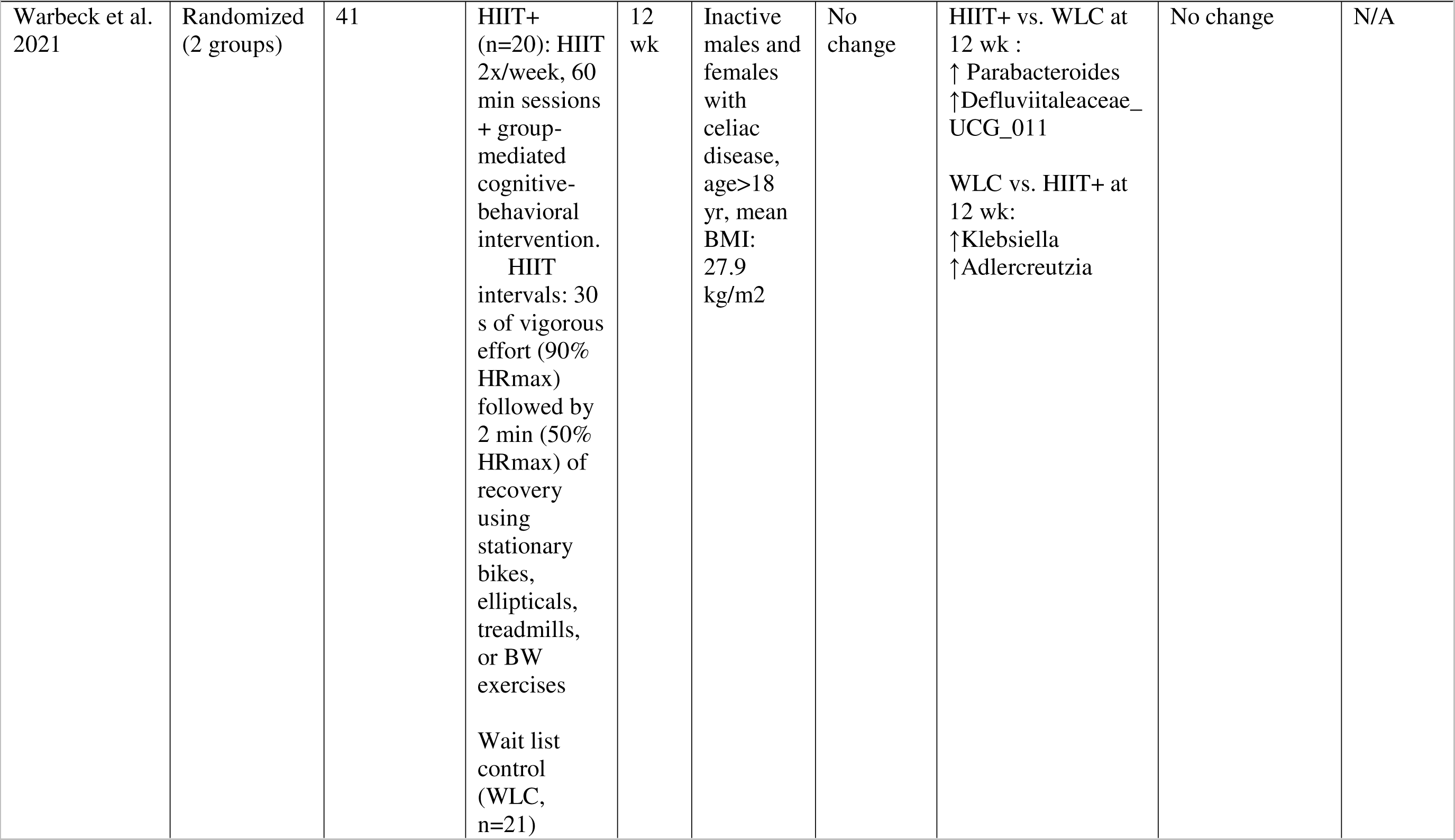
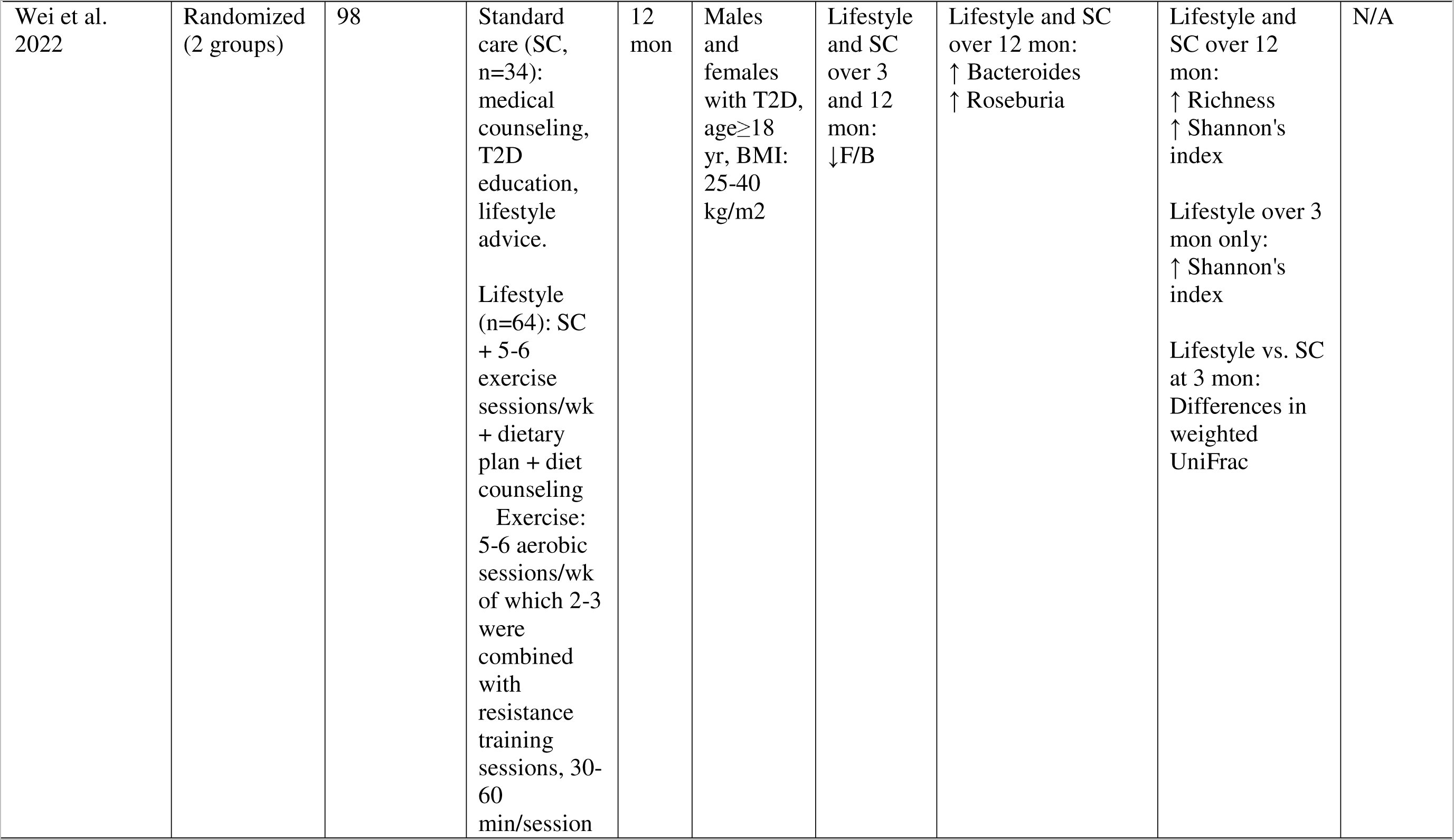
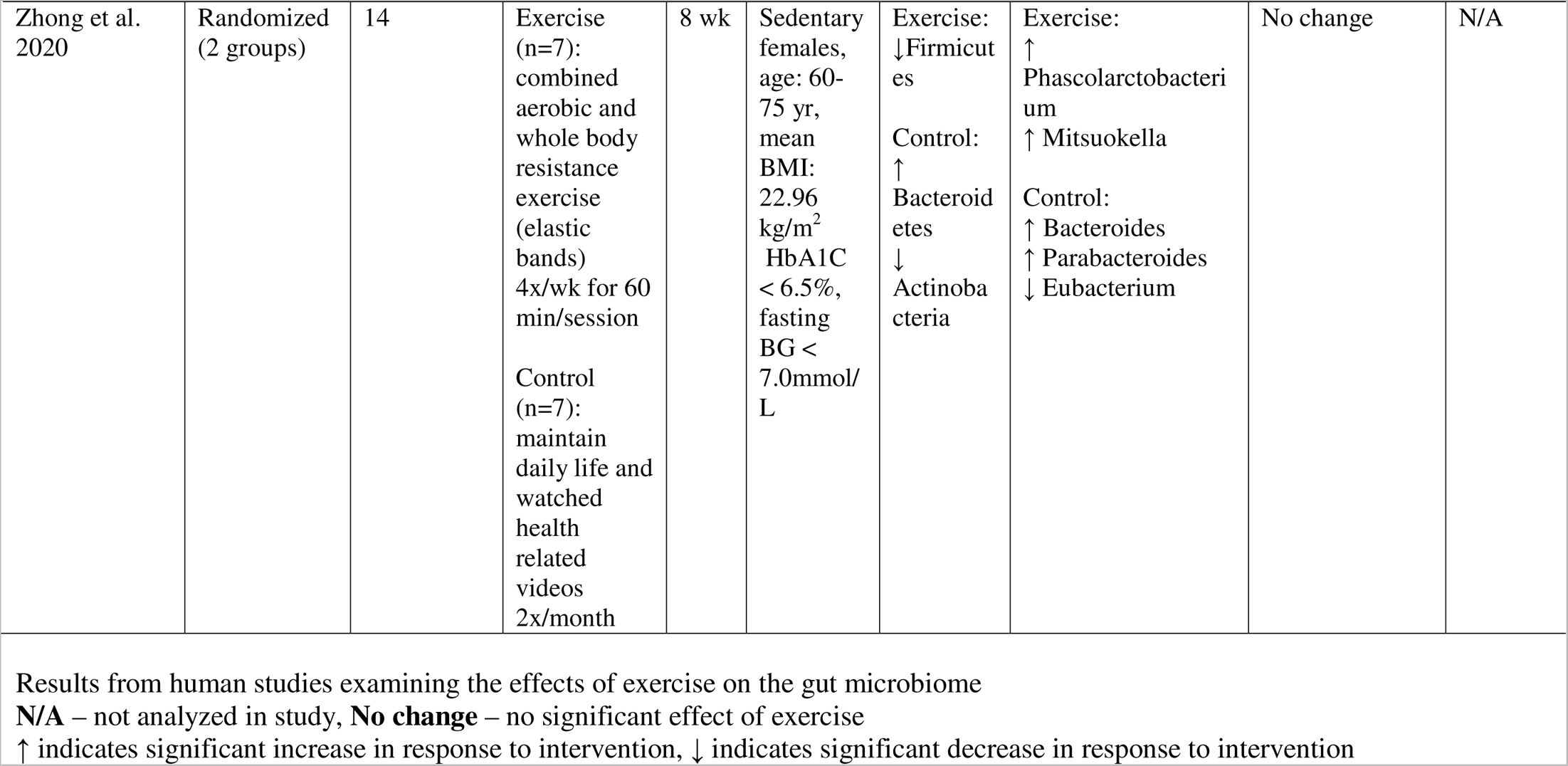
Characteristics of the studies included in the systematic review examining the effects of exercise intervention on gut microbiome outcomes in humans.

Thirteen studies conducted in various animal models met the inclusion criteria for this review (**Table 2**). Of these 13, 5 were conducted in mice (Lamoureux et al., 2017; Houghton et al., 2018; Ribeiro et al., 2019; Chen et al., 2021; Yang et al., 2021a), 4 were in rats (Mika et al., 2015; Giacco et al., 2020; Meng et al., 2020; Castro et al., 2021), and 4 in horses (De Almeida et al., 2016; Janabis et al., 2016; Janabi et al., 2017; Walshe et al., 2021). All but one (Castro et al., 2021) of these studies examined the effects of either aerobic exercise or interventions with aerobic exercise as an important component. Twelve studies were trials with a parallel design (2–8 arms) (Mika et al., 2015; De Almeida et al., 2016; Janabis et al., 2016; Lamoureux et al., 2017; Houghton et al., 2018; Ribeiro et al., 2019; Giacco et al., 2020; Meng et al., 2020; Castro et al., 2021; Chen et al., 2021; Walshe et al., 2021; Yang et al., 2021b), of which the longest trial had a duration of 7 mon (Houghton et al., 2018) and the shortest trials had a duration of 3 days (Giacco et al., 2020). One study followed a crossover design in which horses were exposed to a graded exercise test and a standing control (Janabi et al., 2017). Most mice studies comprised of male and female C57BL/6 mice aged between 3-10 wk. Rats used in the studies were male Wistar rats aged between 6-13 wk (Giacco et al., 2020; Castro et al., 2021), Sprague Dawley rats with obesity aged 16 wk (Meng et al., 2020), and adult male Fischer F344 rats (Mika et al., 2015). Standardbred horses, including mares and geldings, aged 3-17 yr old were the most common horse breed used in the studies.

**Table 2.**
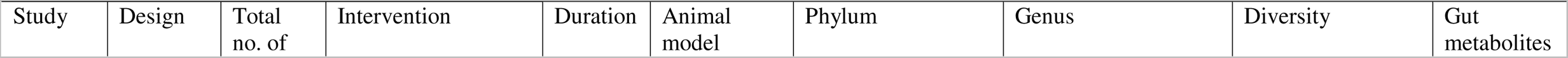
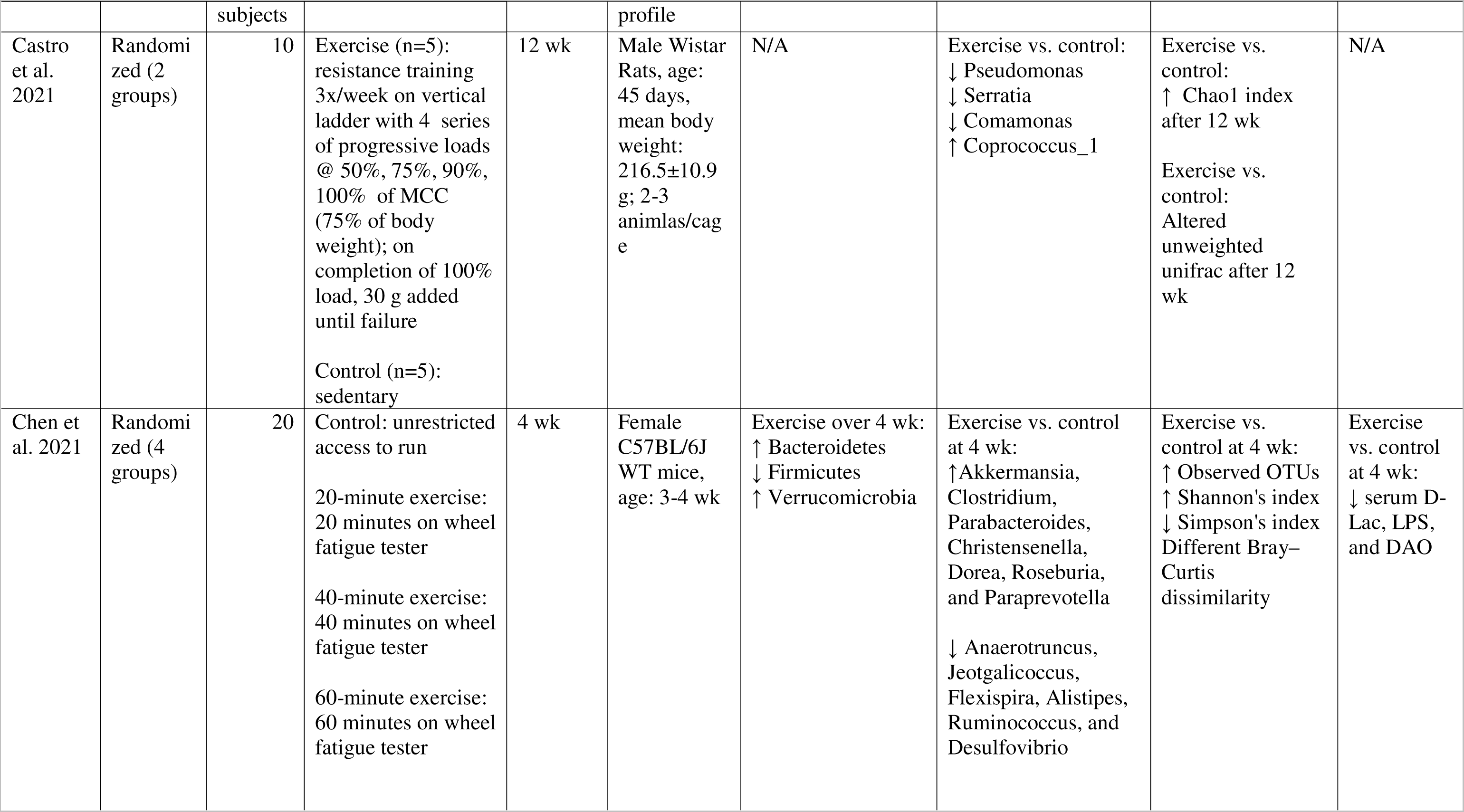
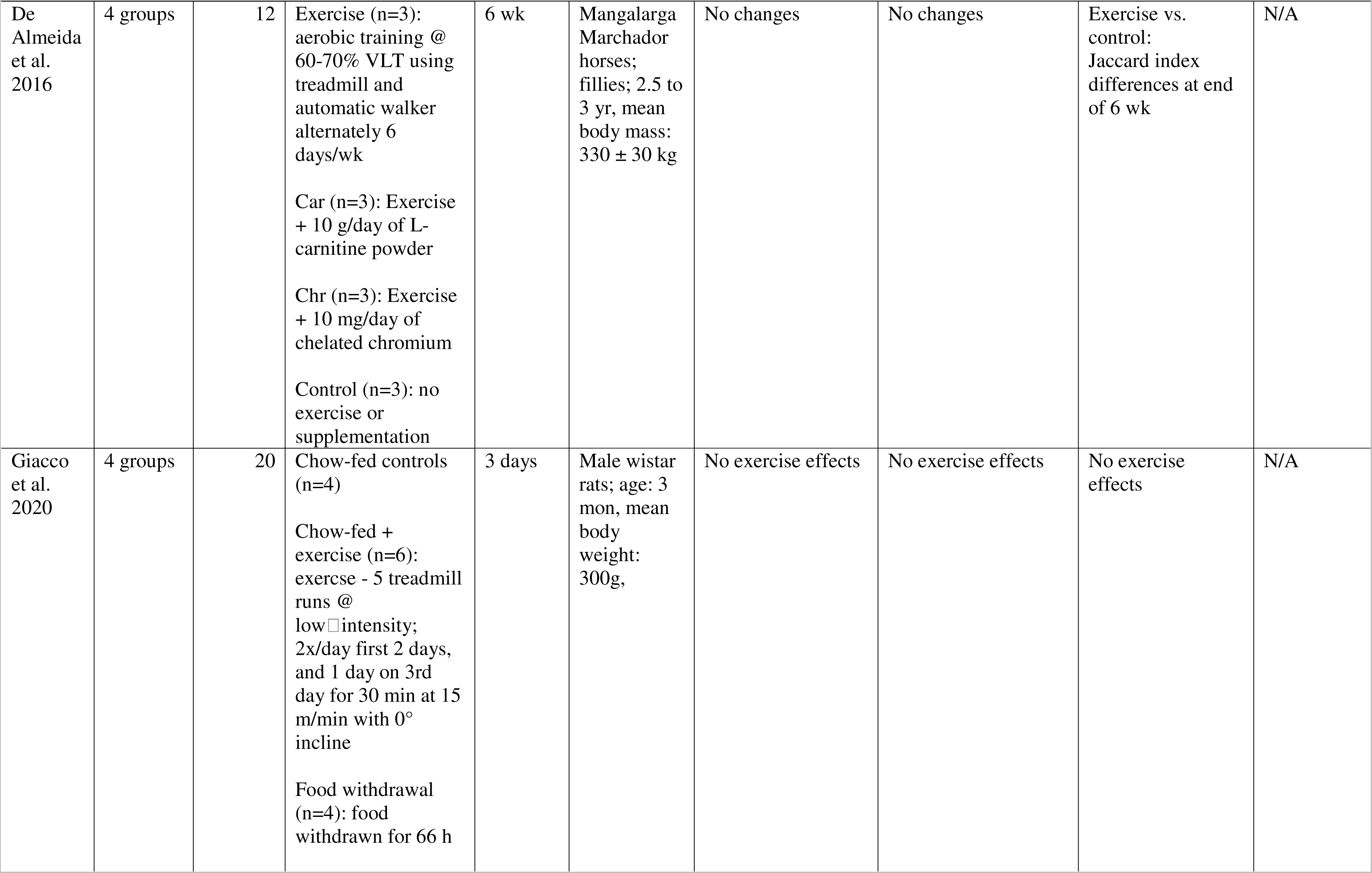
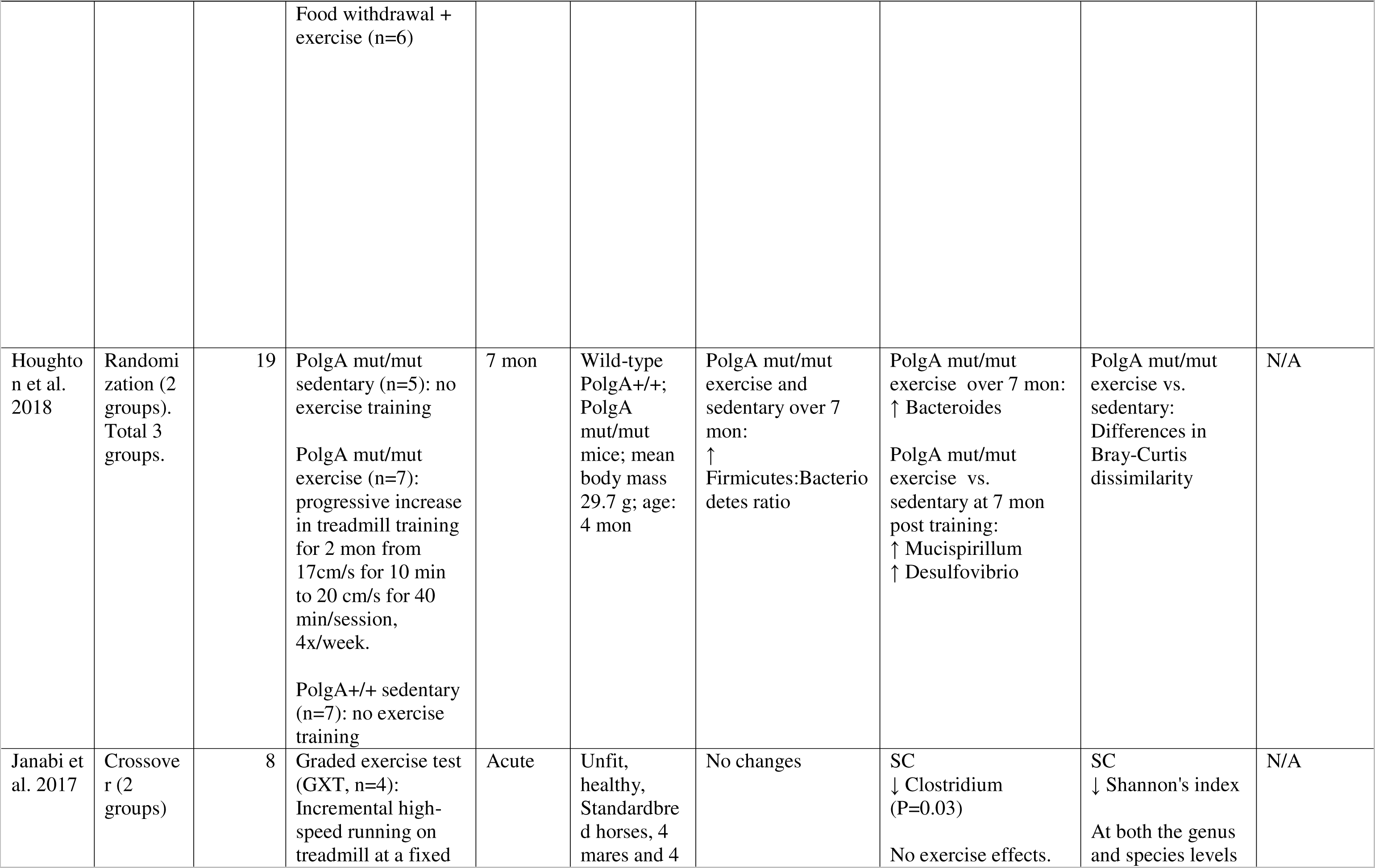
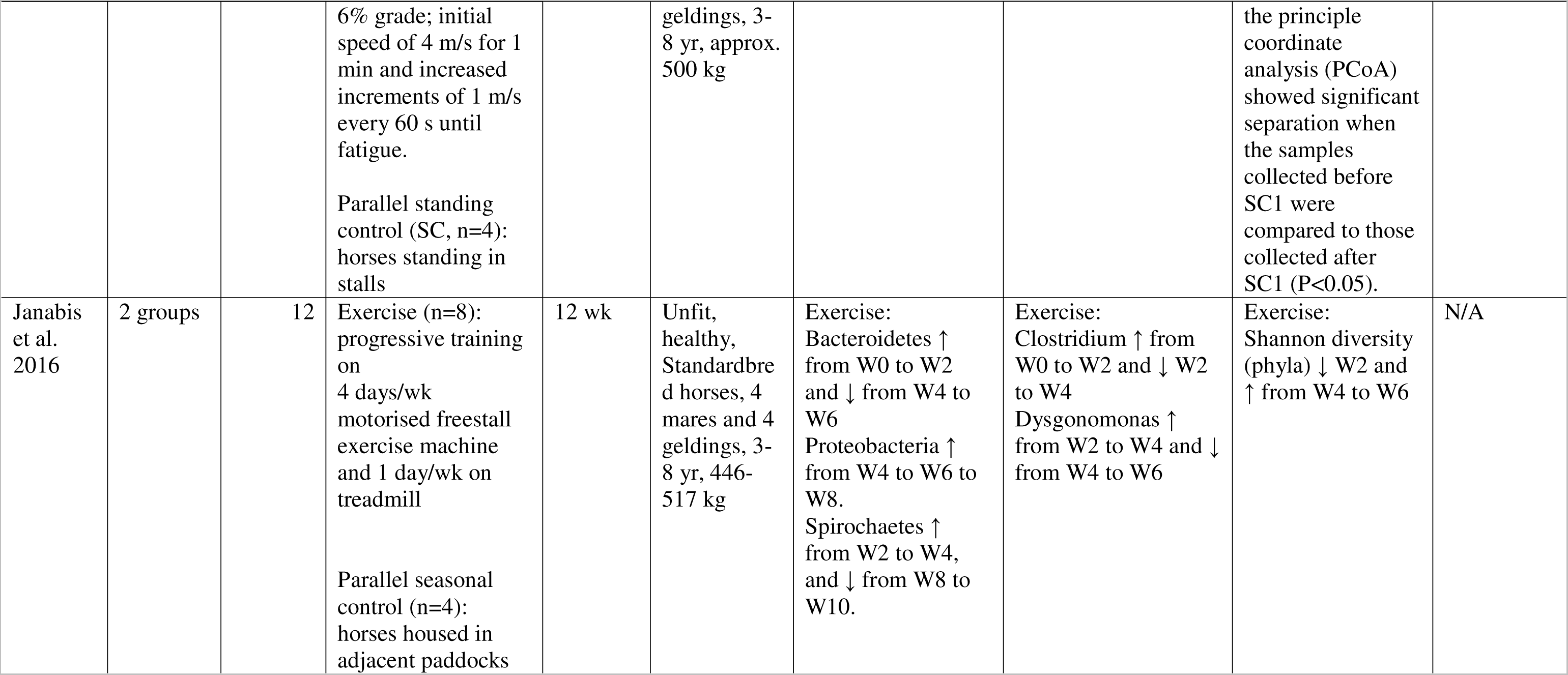
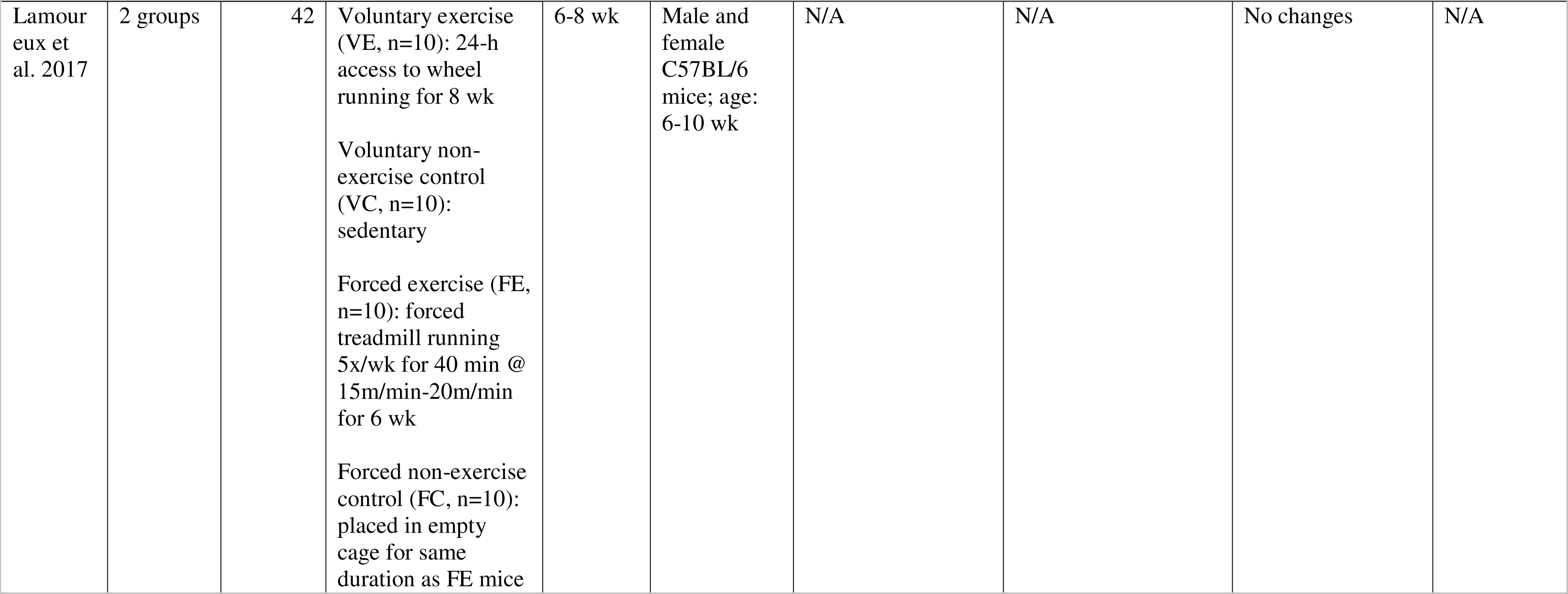
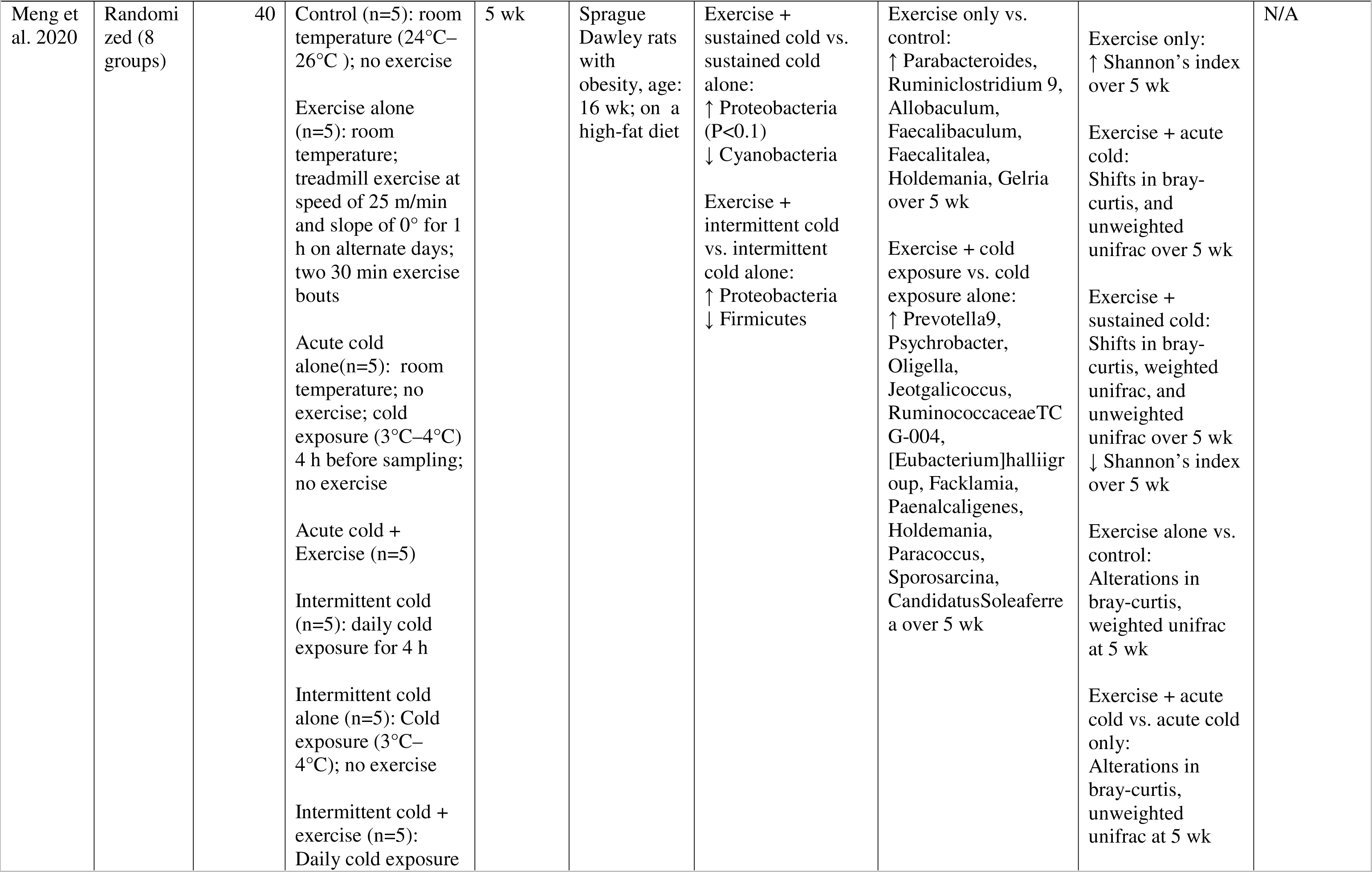
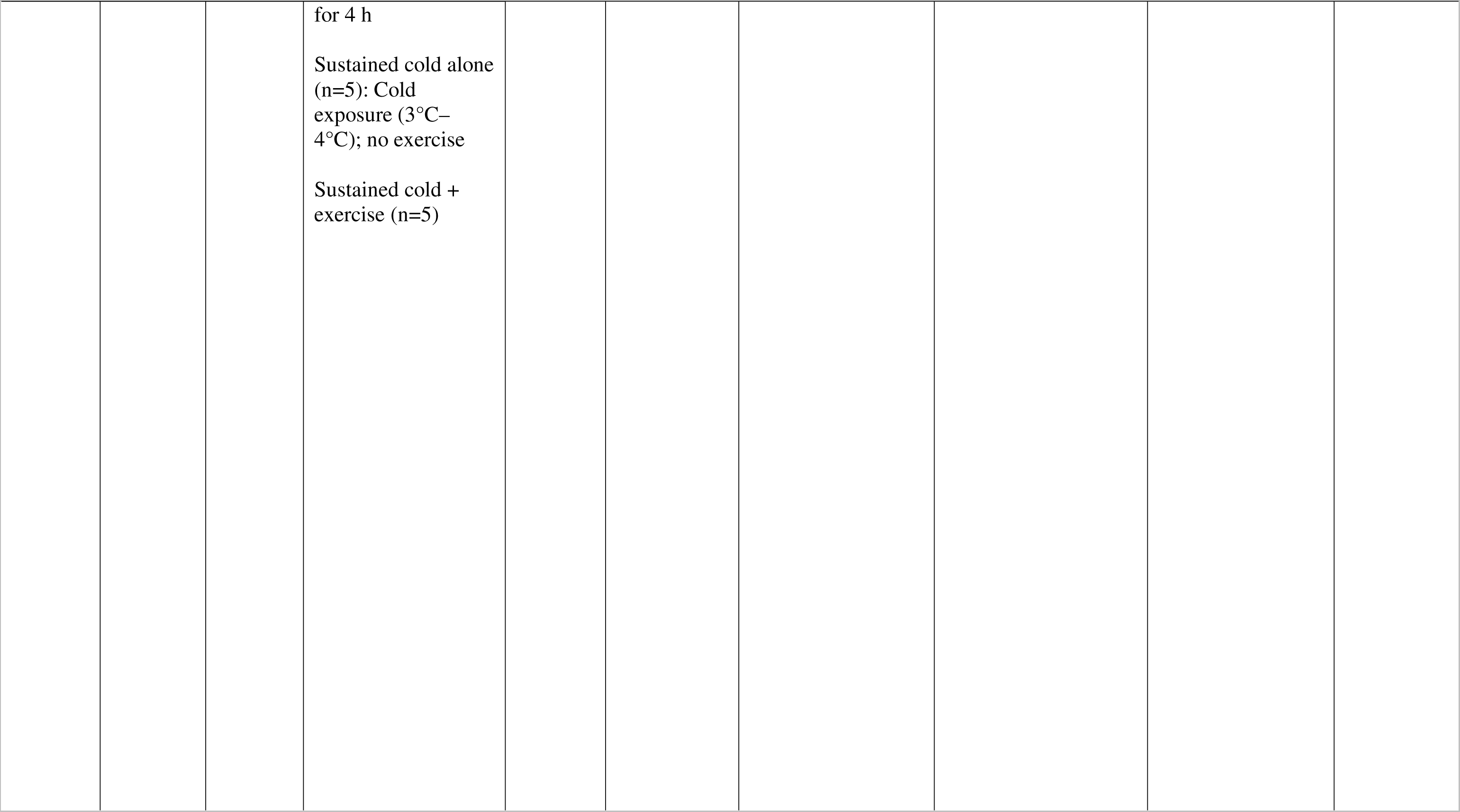
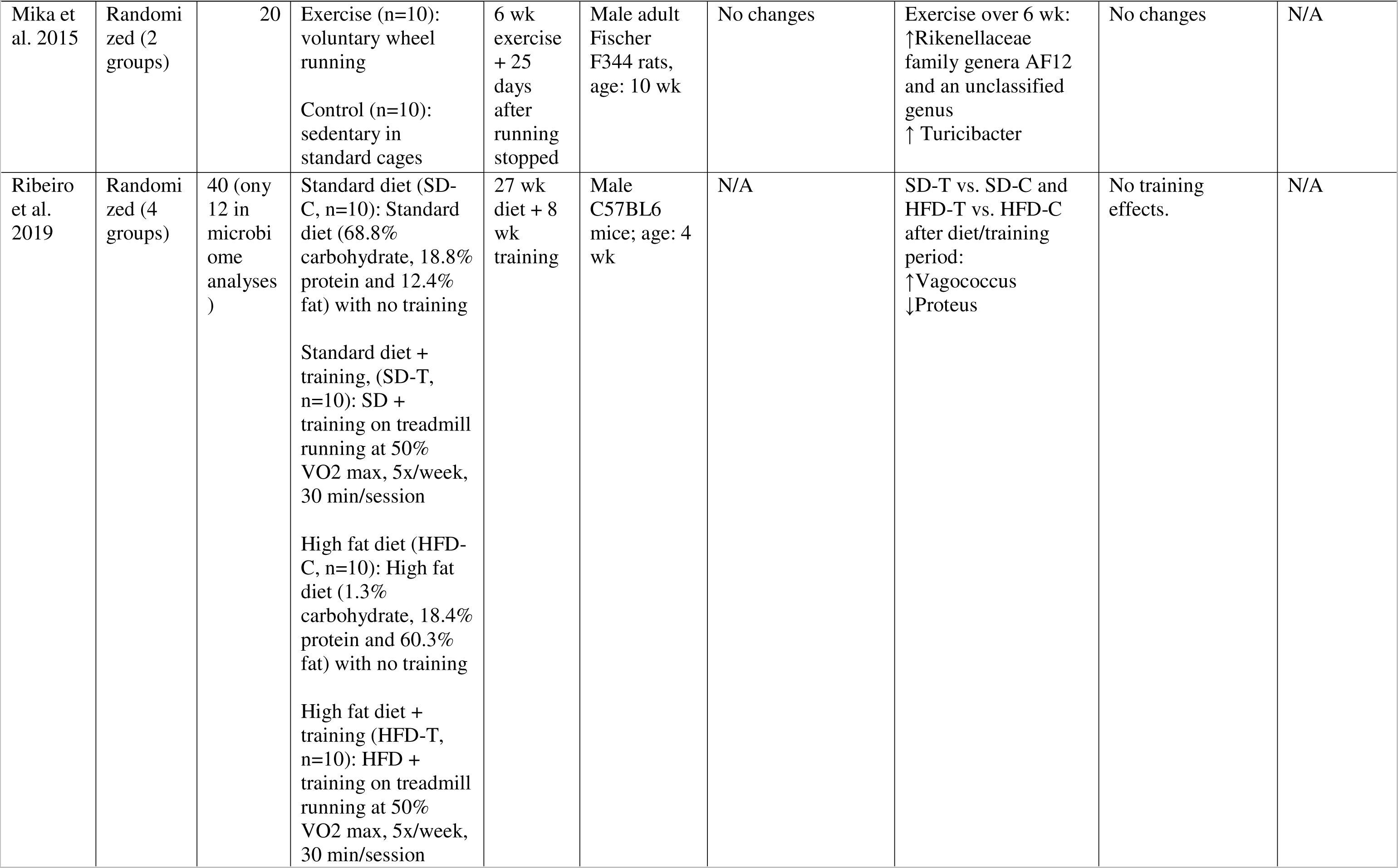
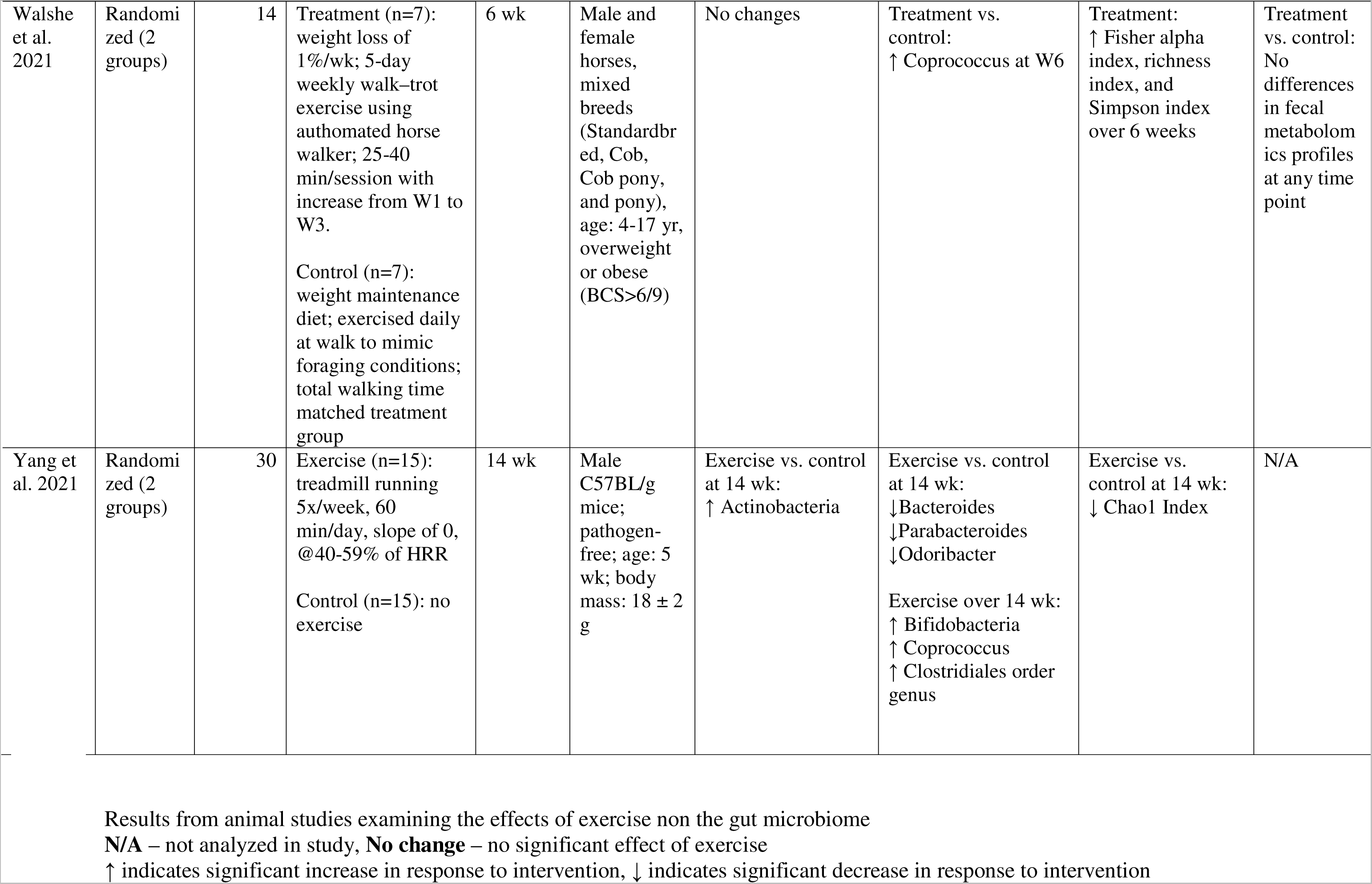
Characteristics of the studies included in the systematic review examining the effects of exercise intervention on gut microbiome outcomes in animal models.

### Effects of exercise interventions on microbial diversity Human studies

Sixteen intervention studies in humans examined microbial diversity outcomes. In a sedentary but healthy population of males and females, 10 wk of aerobic training 3 times a wk, but not resistance training decreased Chao1 index (Moitinho-Silva et al., 2021). In a similar population, six months of moderate and vigorous intensities of leisure time exercise and cycling 5x/wk resulted in a greater shift in Bray-Curtis dissimilarity compared to the non-active control (Kern et al., 2020). Additionally, the cycling group showed a greater change in weighted unifrac diversity while the vigorous intensity group showed an increase in Shannon’s index after 3 months (Kern et al., 2020).

Two studies incorporated exercise with dietary interventions. In one study, combined aerobic and resistance training 3 times a wk for 8 wk along with 24 g protein consumption decreased archeal Shannon’s index but increased bacterial Shannon’s index when compared to the protein only group, and in comparison to the exercise only group, decreased viral Shannon’s index (Cronin et al., 2018a). All three groups showed separation in species Bray-Curtis dissimilarity after the 8 wk intervention (Cronin et al., 2018a). In the other study, a one-year lifestyle intervention resulted in an increase in overall richness and Shannon’s index in individuals with type 2 diabetes which was not different from the standard care control (Wei et al., 2022). The only changes unique to the lifestyle intervention groups were observed differences in weighted unifrac measures at 3 months, however this difference between groups was not maintained throughout the whole intervention (Wei et al., 2022).

In a similar population with diabetes, moderate-intensity continuous training vs. high-intensity exercise interval training (combined aerobic and resistance) resulted in differential effects on Euclidean diversity between the two groups after 8 wk of training (Torquati et al., 2022). In postmenopausal females with overweight and obesity, concurrent HIIT and resistance training for 12 wk resulted in greater change of unweighted unifrac diversity compared to the control (Dupuit et al., 2022). Nine studies demonstrated no change in alpha or beta-diversity with exercise training (Taniguchi et al., 2018; Cronin et al., 2019; Liu et al., 2020; Motiani et al., 2020; Resende et al., 2021; Warbeck et al., 2021; Zhong et al., 2021; Calabrese et al., 2022; Sun et al., 2022).

### Animal studies

Thirteen intervention studies in animal models examined microbial diversity outcomes.

Three mice studies captured differences in diversity outcomes. Aerobic exercise 5x/wk resulted in lower Chao1 index in male B57BL mice when compared to the sedentary control after 14 wk (Yang et al., 2021a). In another study that examined the effects of exercise in response to aging over the course of 7 months found that PolgA mut/mut mice undergoing aerobic exercise 4x/wk had differential effects in Bray-Curtis dissimilarity compared to PolgA mut/mut sedentary at 7 months (Houghton et al., 2018). Aerobic exercise of various durations (20 minutes, 40 minutes, and 60 minutes) resulted in greater observed OTU’s, Shannon’s index, lower Simpson’s index, and differential Bray-Curtis dissimilarity in female C57BL/6J mice at 4 wk compared to control that had unrestricted access to run (Chen et al., 2021).

Two rat studies captured differences in diversity outcomes. In one study that incorporated aerobic exercise at different temperature conditions found that exercise only resulted in increased Shannon’s index over 5 wk, whereas exercise alone vs. control resulted in differential alterations in bray-curtis and weighted unifrac at 5 wk (Meng et al., 2020). A 12 wk resistance training intervention resulted in greater Chao1 index and differential unweighted unifrac diversity in comparison to control after 12 wk (Castro et al., 2021).

Three horse studies captured differences in diversity outcomes. In one study, a twelve-wk aerobic exercise training intervention decreased Shannon’s index at the phyla level after 2 wk of intervention which then increased from wk 4 to wk 6 (Janabis et al., 2016). In another study, aerobic exercise resulted in differential Jaccard diversity at 6 wk in comparison to control (De Almeida et al., 2016). In a weight-loss intervention, aerobic exercise for 6 wk increased Fisher alpha index, richness index, and Simpson index (Walshe et al., 2021).

Five studies demonstrated no change in alpha or beta-diversity with exercise training (Mika et al., 2015; Janabi et al., 2017; Lamoureux et al., 2017; Ribeiro et al., 2019; Giacco et al., 2020).

### Effects of exercise interventions on microbial phyla and genera composition Human studies

Overall, fifteen studies in humans examined taxonomic composition at the phylum and genus levels. Four studies were conducted in healthy individuals. In healthy males, 10 wk of cycling resulted in increased abundance of the genus *Streptococcus* and an unidentified genus in the *Clostridiales* order compared to the non-exercising controls (Resende et al., 2021). In elderly females, brisk walking increased the genus *Bacteroides*, while trunk muscle training demonstrated no effects (Morita et al., 2019). In another study conducted in elderly females, a combined aerobic and resistance training program for 8 wk decreased the phylum *Firmicutes* and increased the genera *Phascolarctobacterium* and *Mitsuokella* (Zhong et al., 2021). In elderly males, a 5 wk cycling intervention increased abundance of the genus *Oscillospira* compared to the control (Taniguchi et al., 2018).

Four studies were conducted in healthy individuals with overweight and obesity. A 10 wk aerobic exercise training intervention in sedentary females increased genus *Bifidobacterium* in comparison to the control (Mahdieh et al., n.d.). In a dietary intervention that incorporated exercise training, the low carbohydrate (LC) diet and aerobic exercise (moderate (MICT) and high intensity (HIIT)) groups exhibited lower abundance of genera *Alistipes* at 4 wk, while the LC-MICT had higher abundance of the genus *Blautia* compared to LC alone (Sun et al., 2022). The LC-HIIT group also decreased *Bifidobacterium* abundance over 4 wk (Sun et al., 2022). Two studies did not find any effects of exercise training on phyla or genera composition (Kern et al., 2020; Dupuit et al., 2022).

Seven studies were conducted in populations with pre-diabetes, diabetes, NAFLD, IBD, Multiple Sclerosis (MS), or celiac disease.

In a T2D population, moderate-intensity combined aerobic and resistance training led to higher collective abundance of phyla *Verrucomicrobia, Actinobacteria,* and *Desulfobacterota,* and genus *Bifidobacterium* compared to high-intensity combined aerobic and resistance training after 8 wk (Torquati et al., 2022). In a prediabetic and diabetic population, both moderate intensity continuous training and sprint interval training for 2 wk decreased the Firmicutes/Bacteroidetes ratio driven by an increase in Bacteroidetes, and decreased genera *Blautia* and *Closridium*, while only the moderate continuous training group increased genera *Veillonella* and *Faecalibacterium*, and only sprint interval training increased genus *Lachnospira* (Exercise Training Modulates Gut Microbiota Profile and Improves Endotoxemia..pdf, n.d.). In a diabetic population, a 12-month lifestyle intervention consisting of dietary counseling and exercise decreased the Firmicutes/Bacteroidetes ratio and increased abundance of genera *Bacteroides* and *Roseburia* similar to the standard of care (Wei et al., 2022).

In individuals with NAFLD, the aerobic exercise only group exhibited greater genera *Bilophila, Roseburia, Erysipelotrichaceae_UCG_003, Hungatella, and Lachnospiraceae_UCG_004* whereas the aerobic exercise plus diet group exhibited greater genera *Alistipes*, *Bacteroides*, *Bilophilla, Butyricimonas, Roseburia,* and several genera in the *Lachnospiriceae and Ruminococcaceae* families (Cheng et al., 2022) after the intervention in comparison to the control. In another study, a low glycemic index Mediterranean diet in conjunction with an aerobic exercise intervention for 3 months led to higher abundance of genera *Ruminococcus, Lachnospiraceae_GCA900066575, Clostridia VadinBB60 group, Enterorabdus, Coprobacter, UCG002 (Oscillospiraceae), Intestinimonas, and Ruminococcaceae_g_UBA1819*, and lower abundance of *Coprococcus* in individuals with NAFLD in comparison to the diet and exercise alone groups (Calabrese et al., 2022).

In individuals with MS, a 6-month home-based combined aerobic and exercise training program increased genus *Prevotella* compared to a non-exercising control (Mokhtarzade et al., 2021). In individuals with celiac disease, a 12 wk HIIT intervention enriched genera *Parabacteroides* and *Defluviitaleaceae* (Warbeck et al., 2021).

### Animal studies

Twelve intervention studies in animal models examined taxonomic composition at the phylum or genus levels.

All four studies in mouse models examined the effects of aerobic exercise on the gut microbiome. Aerobic exercise 5x/wk resulted in higher abundance of phylum *Actinobacteria* and lower abundance of genera *Bacteroides, Parabacteroides,* and *Odoribacteria* in male B57BL mice compared to control group after 14 wk (Yang et al., 2021a). Moreover, the exercise group showed an increase in the genera *Bifidobacterium*, *Coprococcus*, and an unidentified genus in the *Clostridiales* order over 14 wk (Yang et al., 2021a). In a similar model of male B57BL mice, the effects of a high fat diet were compared to a standard chow diet with and without 8 wk of aerobic exercise (Ribeiro et al., 2019). They only observed changes at the genus level with both exercise groups exhibiting higher *Vagococcus* and lower *Proteus* abundances after the training period when compared to their non-exercising controls. In another study, aerobic exercise led to higher phyla *Bacteroidetes* and *Verrucomicrobia* and lower *Firmicutes* compared to the control at 4 wk (Chen et al., 2021). At the genus level, exercise led to higher *Akkermansia, Clostridium, Parabacteroides, Christensenella, Dorea, Roseburia*, and *Paraprevotella* and lower *Anaerotruncus, Jeotgalicoccus, Flexispira, Alistipes, Ruminococcus*, and *Desulfovibrio* compared to the control at 4 wk (Chen et al., 2021). In the PolgA mut/mut mice study (Houghton et al., 2018), aerobic exercise for 7 months increased the Firmicutes:Bacteriodetes ratio and the genus *Bacteroides.* Additionally, the exercise groups exhibited a higher abundance of the genera *Muucispirillum* and *Desulfovibrio* when compared to the non-exercised controls at 7 months (Houghton et al., 2018).

Four studies conducted in rats examined the effects of exercise training on phyla or genera outcomes. A 12 wk resistance training intervention in male Wistar rats resulted in decreased *Pseudomonas*, *Serratia*, *Comamonas* but increased *Coprococcus_1* in comparison to the control (Castro et al., 2021). In the study that incorporated aerobic exercise at different temperature conditions, exercise at room temperature for 5 wk resulted in increases in the genera *Parabacteroides*, *Ruminiclostridium*, *Allobaculum*, *Faecalibaculum*, *Faecalitalea*, *Holdemania*, and *Gelria* when compared to the non-exercising control (Meng et al., 2020). Exercise with different modalities of cold exposure also resulted in other changes at the phylum and genera level which were not consistent for exercise as whole (Meng et al., 2020). Moreover, in a voluntary wheel running study, exercise for 6 wk resulted in increased *Rikenellaceae* family genera *AF12* and an unclassified genus, and *Turicibacter* (Mika et al., 2015). One rat study did not find any effect of exercise training on phylum or genus taxonomic composition (Giacco et al., 2020).

Four studies conducted in horse models tested the effects of aerobic exercise and/or other lifestyle interventions on phyla or genera outcomes. In a weight-loss intervention, aerobic exercise led to higher abundance of the genus *Coprococcus* compared to the non-exercising control group at 6 wk (Walshe et al., 2021). In another study, a 12 wk aerobic exercise intervention led to an increase of the phylum *Bacteroidetes* in the first 2 wk and then a decrease from wk 4 to wk 6 (Janabis et al., 2016). A progressive increase in the phylum *Proteobacteria* was also observed from wk 4 to wk 6 to wk 8, and in the phylum *Spirochaetes* from wk 2 to wk 4, however there was a reduction in *Spirochaetes* from wk 8 to wk 10 in response to exercise. At the genus level, *Clostridium* increased from wk 0 to wk 2 and decreased from wk 2 to wk 4, whereas *Dysgonomonas* increased from wk 2 to wk 4 and decreased from wk 4 to wk 6 in response to exercise. Two horse studies did not find any effect of exercise training on phyla or genus taxonomic composition (De Almeida et al., 2016; Janabi et al., 2017).

### Effects of exercise interventions on microbial metabolites

#### Human studies

Only three human studies assessed effects of exercise training on gut microbiome derived metabolites. High-intensity combined aerobic and resistance training for 12 wk in males with overweight and obesity resulted in a decrease in fecal and serum BCAA and aromatic AA, and an increase in fecal propionate and GABA, and serum SCFA in exercise responders (Liu et al., 2020). Additionally, Cronin et al. (Cronin et al., 2018a) found that the a combined aerobic resistance training program of moderate intensity for 8 wk resulted in a greater decrease in urinary PAG and TMAO compared to the protein only group. A similar study did not find any effects of exercise training on fecal SCFAs in individuals with type 2 diabetes (Torquati et al., 2022).

#### Animal studies

Only two animal studies assessed gut microbial metabolites. Four wk of aerobic training of different durations (20,40, or 60 min) in female C57BL/6J WT mice resulted in lower serum D-Lac, LPS, and DAO in comparison to unrestricted running control (Chen et al., 2021). However, no differences in fecal metabolomics profiles were found with aerobic training in overweight or obese mixed breed horses undergoing weight-loss (Walshe et al., 2021).

### Risk of bias and quality of assessment of studies included in systematic review

Characteristics of the included human studies are summarized in **Table 3**. Seven articles were rated positive, indicating that they had adequately addressed issues of bias, generalizability, and data collection. The remaining 12 were found to be neutral, meaning they were neither exceptionally weak nor exceptionally strong. Characteristics of the included animal studies are summarized in **Table 4**. One article was rated as positive, 11 as neutral, and 1 as negative, indicating that it has inadequately addressed validity criteria. Overall, the human and animal studies lacked consistency in reporting of statistical analyses and interpretation of data. The issues with statistical analyses pertain to not reporting or including overall exercise group by time interaction effects, or adjusting post-intervention values for pre-intervention values in the statistical model, not accounting for repeated measures, restricting analysis to only selected exercise arms without specifying a priori hypotheses, insufficient description of the statistical model for interpretation of results, or interpretation based on post hoc tests even when interaction effects were not significant.

**Table 3.**
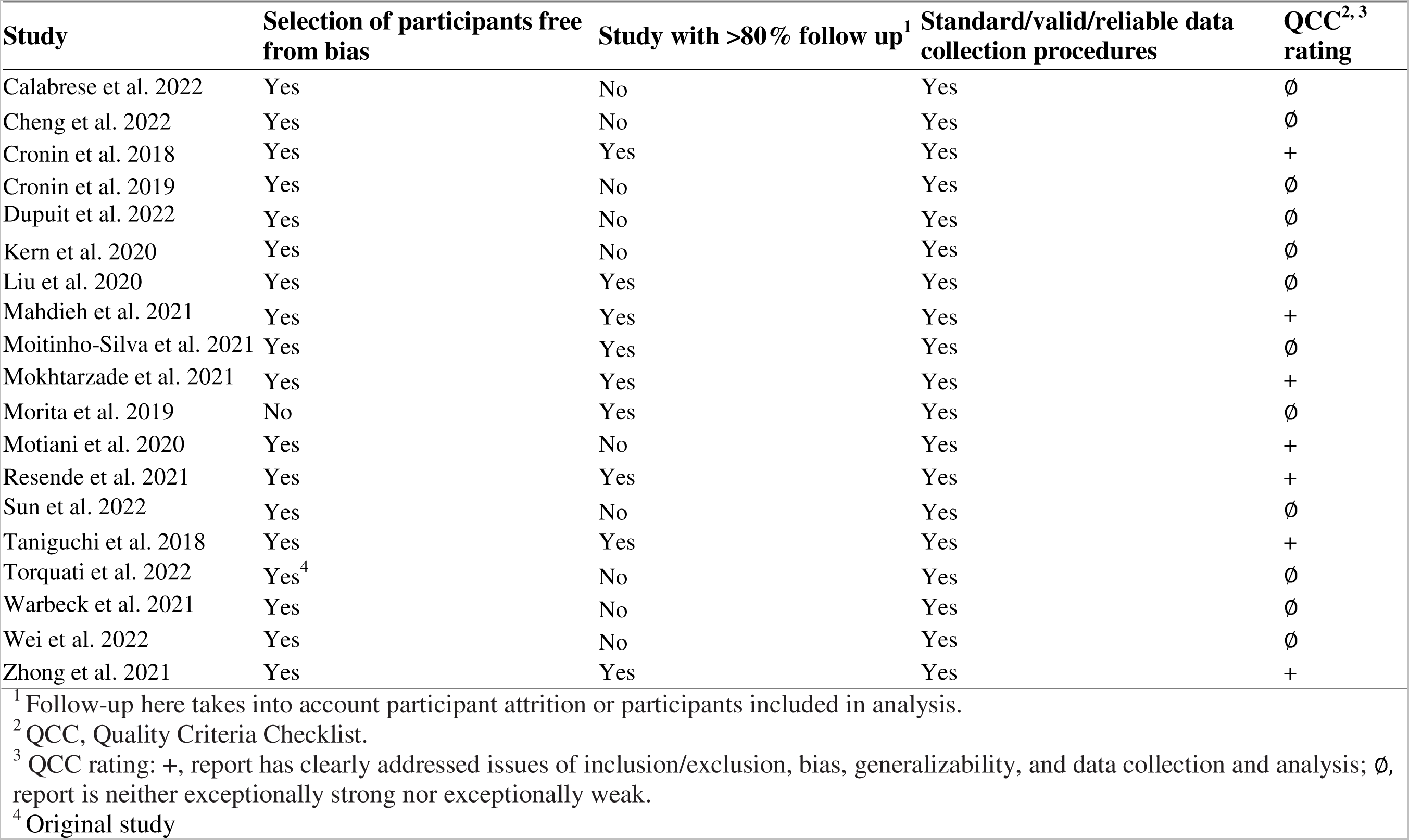
Risk-of-bias analysis of studies included in the systematic review examining the effects of exercise interventions on the gut microbiome in humans.

**Table 4.** Risk-of-bias analysis of studies included in the systematic review examining the effects of exercise interventions on the gut microbiome in animal models.

| Study | Selection of participants<br>free from bias. | Study with >80% follow<br>up<sup>1</sup> | Standard/valid/reliable data<br>collection procedures | QCC<br>rating<sup>2,3</sup> |
| --- | --- | --- | --- | --- |
| Castro et al. 2021 | Yes | Yes | Yes | Ø |
| Chen et al. 2021 | Yes | Unclear | Yes | - |
| De Almeida et al. 2016 | Yes | Yes | Yes | + |
| Giacco et al. 2020 | Yes | No | Yes | Ø |
| Houghton et al. 2018 | Yes | Unclear | Yes | Ø |
| Janabi et al. 2017 | Yes | Yes | Yes | Ø |
| Janabis et al. 2016 | Yes | Yes | Yes | Ø |
| Lamoureux et al. 2017 | Yes | Unclear | Yes | Ø |
| Meng et al. 2020 | Yes | Yes | Yes | Ø |
| Mika et al. 2015 | Yes | No | Yes | Ø |
| Ribeiro et al. 2019 | Yes | No | Yes | Ø |
| Walshe et al. 2021 | No<sup>4</sup> | Yes | Yes | Ø |
| Yang et al. 2021 | Yes | Unclear | Yes | Ø |
| 775 | <sup>1</sup> Follow-up here takes into account participant attrition or participants included in analysis. |  |  |  |
| 776 | <sup>2</sup> QCC, Quality Criteria Checklist. |  |  |  |
| 777 | <sup>3</sup> QCC rating: +, report has clearly addressed issues of inclusion/exclusion, bias, generalizability, and data collection and analysis; -, |  |  |  |
| 778 | report has not addressed these issues adequately; and Ø, report is neither exceptionally strong nor exceptionally weak. |  |  |  |
| 779 | <sup>4</sup> Client-owned, not experimental animals |  |  |  |

## 4. Discussion

### Summary of evidence

More than 50% of the studies did not demonstrate any exercise effects on microbial diversity in humans. Of those that did, exercise was found to generally favor an increase in Shannon’s index, an index of microbial richness and evenness, regardless of disease status. However, when combined with dietary changes, the impact on alpha-diversity can vary. For instance, one study demonstrated opposite effects for archaeal versus bacterial Shannon’s index in response to a combined aerobic and dietary protein intervention (Cronin et al., 2018b). The weighted unifrac measure, a phylogenetic measure that accounts for the relatedness of microbial taxa, was the most commonly reported beta-diversity metric. Changes in other beta-diversity measures, like Bray-Curtis dissimilarity, both weighted and unweighted Unifrac, and Euclidean diversity, were also documented regardless of disease status and exercise type. The direction of these changes, however, wasn’t specified. A greater proportion of animal studies reported changes in diversity compared to human studies. However, no clear pattern emerged due to the mixed effects of exercise (predominantly aerobic) on alpha or beta-diversity metrics.

According to ecological theory, diversity in the gut microbiome, both in terms of species richness and functional response, is integral to its resilience (Lozupone et al., 2012). A diverse microbiome is better equipped to resist invaders and efficiently use resources. Notably, even when certain species are compromised, others, with similar functions, can step in, ensuring stability. This phenomenon is further demonstrated by the functional redundancy seen in various bacteria in the gut. Thus, a richer and functionally diverse microbiome may offer greater stability and adaptability in the face of disturbances (Lozupone et al., 2012). Hence, the success of an exercise intervention may be predicated upon the baseline microbiome composition of individuals, which in turn can be influenced by disease status, dietary habits, and other environmental factors. Whether individuals with a less diverse or less stable microbiome at baseline may be more susceptible to changes in microbial composition with exercise compared to individuals with a more diverse and stable microbiome is a hypothesis that should be tested further. And even if overall diversity doesn’t change significantly with exercise, individual taxa can still be impacted in terms of their relative abundance or metabolic activity.

Our review indicates that exercise interventions can also affect taxonomic composition at the phylum and genus levels. In humans, at the phyla level, exercise was typically associated with a decrease in the Firmicutes/Bacteroidetes ratio. At the genera level, while *Bacteroides*, *Roseburia*, *Parabacteroides*, *Blautia*, *Alistipes* were most reported to change in response to exercise in humans, only *Bacteroides* and *Roseburia*, demonstrated consistent increases with exercise. Bacteroides is a genus involved in many metabolic pathways including sphingolipid production (Johnson et al., 2020b), SCFA production, bile acid metabolism (Ghosh et al., 2021), and mucus utilization (Wang et al., 2023). *Roseburia* is majorly involved in the production of SCFAs, particularly butyrate, and in conjugated fatty acid metabolism (Ghosh et al., 2021). Both SCFA and conjugated fatty acids, play a role in regulation of intestinal permeability, while SCFA are also associated with energy homeostasis, and increased glucose tolerance and insulin sensitivity. Amongst the other genera that showed no consistent effects with exercise, *Blautia* has been associated with potential probiotic capabilities but is also enriched in IBD (Liu et al., n.d.), *Alistipes* has been found to be pathogenic in colorectal cancer but has protective effects in CVD (Parker et al., 2020), while *Parabacteroides* is implicated in colitis (Dziarski et al., 2016) but also demonstrates a protective effect in obesity (Wang et al., 2019).

In animal models collectively, *Parabacteroides, Coprococcus*, *Bacteroides*, and *Clostridium* were most reported to change in response to exercise. However, only *Coproccous*, another SCFA producer, demonstrated consistent increases with exercise. In general, in both human and animal models, the diversity in intervention duration, exercise modality and intensity, disease status, dietary factors, baseline microbiota composition etc. appear to have a huge effect on individual microbial taxa, and due to the limited number of studies, stratification by these factors was not possible. A common pattern though was an exercise-induced increase in SCFA producers such as *Christensenella*, *Dorea*, *Roseburia*, *Paraprevotella*, *Ruminiclostridium*, and *Allobaculum*. Our review of the literature also identified SCFA as the most measured metabolite. However, due to the limited human and animal studies examining exercise effects on microbial-produced metabolites including SCFAs, no clear patterns emerged.

### Strengths and limitations of review

While a few systematic reviews on this topic have been published (Mailing et al., 2019; Mitchell et al., 2019; Tzemah Shahar et al., 2020; Clemente et al., 2021; Hughes and Holscher, 2021), the uniqueness of this systematic review lies in the inclusion of only studies that have a control or other arm for rigorous comparisons to the intervention arms, and inclusion of both human and animal studies for a comprehensive overview of the exercise-microbiome literature. However, like any field in the initial stages, the heterogeneity of exercise intervention designs, taxa outcomes profiled, and populations recruited, limited the ability to make direct comparisons among studies. Moreover, several studies included in this review did not account for confounding factors such as dietary habits, medication use, or other environmental factors, that could influence the relationship between exercise and the microbiome.

### Implication of findings, research gaps and future directions

This research underscores the importance of the gut microbiome in personalized health and precision medicine by providing insights into the potential benefits of exercise on gut microbial diversity and composition. However, the lack of standardized exercise protocols across studies makes it challenging to compare results and draw conclusive findings. Establishing standardized protocols would enable better comparisons and generalizability of results. Additionally, there is a need for adequately powered studies and rigorous statistical models to accurately interpret group by time effects in longitudinal studies. In the context of precision medicine, considering factors such as disease status, exercise modality/intensity, and baseline microbiota composition can optimize the selection and customization of exercise interventions, leading to more effective therapeutic strategies for improving gut health.

An important research gap in the exercise-microbiome field is the limited utilization of interspersed designs that incorporate baseline microbiome composition. Currently, the typical practice is to employ randomization without considering the initial microbial profiles of participants. However, accounting for baseline microbiome composition or stratifying participants based on baseline microbiome composition ranges would capture a broader range of responses to exercise interventions and allow for assessing how these responses depend on the presence of selected microbes (Johnson et al., 2020a). Understanding an individual’s baseline microbiome composition and its response to exercise is crucial for personalized health. Individuals with a less diverse or stable microbiome at baseline may be more susceptible to changes in microbial composition with exercise, emphasizing the importance of tailored interventions to optimize gut health benefits. Implementing such designs would also improve statistical power.

Furthermore, the identification of specific genera, such as *Bacteroides*, *Roseburia*, and *Coprococcus*, as consistent responders to exercise highlights their potential as targets for interventions and biomarkers of exercise-induced changes in the gut microbiome. Future research should focus on elucidating the underlying mechanisms of these changes and their impact on health outcomes. While changes in microbial composition have been widely studied, investigating the specific microbial metabolites influenced by exercise is crucial to unravel the functional implications of exercise-induced microbiome changes. To gain a better understanding of the variability in responses and potential mechanistic insights, it is important to consider various factors such as employing ‘omics techniques to examine biological factors and taking into account the socioecological context. Other key considerations in microbiome research from other reviews (Knight et al., 2018; Bharti and Grimm, 2021; Shanahan et al., 2021; Mattes et al., 2022), although mostly for diet studies, are also applicable to exercise studies.

### Conclusions

In conclusion, this systematic review reveals that exercise has the potential to influence the gut microbiome, with indications of increased microbial diversity and changes in taxonomic composition. Further research is needed to standardize exercise protocols, incorporate larger sample sizes, and employ rigorous statistical models. Additionally, investigating the functional implications of exercise-induced changes in microbial metabolites and understanding the underlying mechanisms will enhance our understanding of the exercise-microbiome relationship. Considering the growing prevalence of chronic diseases, such as obesity, diabetes, and cardiovascular disorders, studying the exercise-microbiome relationship may provide valuable insights for preventive and therapeutic interventions. Targeted exercise programs that promote a favorable microbial profile could potentially serve as a non-invasive and cost-effective approach to support overall health and mitigate the risk of chronic diseases. These advancements can guide the development of personalized exercise interventions for optimizing health.

## Conflict of Interest

The authors declare that the research was conducted in the absence of any commercial or financial relationships that could be construed as a potential conflict of interest.

## Author contributions

The authors’ responsibilities were as follows—JD and JC: conceptualized the study; JD, JC, and SS: extracted and analyzed data; JD, JC, and SS: wrote the manuscript; and all authors read and approved the final manuscript and take responsibility for the final content.

## Data Availability

Not applicable

## Notes

### Competing Interest Statement

The authors have declared no competing interest.

### Funding Statement

This study did not receive any funding

## References

Academy of Nutrition and Dietetics (n.d.). Evidence Analysis Manual. Available at: https://www.andeal.org/evidence-analysis-manual [Accessed April 5, 2023].

Barton, W., Penney, N. C., Cronin, O., Garcia-Perez, I., Molloy, M. G., Holmes, E., et al. (2018). The microbiome of professional athletes differs from that of more sedentary subjects in composition and particularly at the functional metabolic level. Gut 67, 625–633. doi: 10.1136/gutjnl-2016-313627.

Bharti, R., and Grimm, D. G. (2021). Current challenges and best-practice protocols for microbiome analysis. Brief Bioinform 22, 178–193. doi: 10.1093/bib/bbz155.

Calabrese, F. M., Disciglio, V., Franco, I., Sorino, P., Bonfiglio, C., Bianco, A., et al. (2022). A Low Glycemic Index Mediterranean Diet Combined with Aerobic Physical Activity Rearranges the Gut Microbiota Signature in NAFLD Patients. Nutrients 14. doi: 10.3390/nu14091773.

Castro, A., Silva, K., Medeiros, C., Alves, F., Araujo, R., and Almeida, J. (2021). Effects of 12 weeks of resistance training on rat gut microbiota composition. JOURNAL OF EXPERIMENTAL BIOLOGY 224. doi: 10.1242/jeb.242543.

Chen, H., Shen, L., Liu, Y., Ma, X., Long, L., Ma, X., et al. (2021). Strength Exercise Confers Protection in Central Nervous System Autoimmunity by Altering the Gut Microbiota. FRONTIERS IN IMMUNOLOGY 12. doi: 10.3389/fimmu.2021.628629.

Cheng, R., Wang, L., Le, S., Yang, Y., Zhao, C., Zhang, X., et al. (2022). A randomized controlled trial for response of microbiome network to exercise and diet intervention in patients with nonalcoholic fatty liver disease. Nature Communications 13. doi: 10.1038/s41467-022-29968-0.

Cheng, Y., Xie, G., Chen, T., Qiu, Y., Zou, X., Zheng, M., et al. (2012). Distinct urinary metabolic profile of human colorectal cancer. Journal of Proteome Research 11, 1354– 1363. doi: 10.1021/pr201001a.

Clarke, S. F., Murphy, E. F., O’Sullivan, O., Lucey, A. J., Humphreys, M., Hogan, A., et al. (2014). Exercise and associated dietary extremes impact on gut microbial diversity. Gut 63, 1913–1920. doi: 10.1136/gutjnl-2013-306541.

Clemente, F., Bravini, E., Corna, S., Colombo, E., Sartorio, F., Rinaldi, C., et al. (2021). [The relationship between physical exercise and gut microbiota in the human being: a systematic review]. Epidemiol Prev 45, 245–253. doi: 10.19191/EP21.4.P245.080.

Cochrane Handbook for Systematic Reviews of Interventions (n.d.). Available at: https://training.cochrane.org/handbook [Accessed January 12, 2023].

Cronin, O., Barton, W., Moran, C., Sheehan, D., Whiston, R., Nugent, H., et al. (2019). Moderate-intensity aerobic and resistance exercise is safe and favorably influences body composition in patients with quiescent Inflammatory Bowel Disease: A randomized controlled cross-over trial. BMC Gastroenterology 19. doi: 10.1186/s12876-019-0952-x.

Cronin, O., Barton, W., Skuse, P., Penney, N. C., Garcia-Perez, I., Murphy, E. F., et al. (2018a). A prospective metagenomic and metabolomic analysis of the impact of exercise and/or whey protein supplementation on the gut microbiome of sedentary adults. mSystems 3. doi: 10.1128/mSystems.00044-18.

Cronin, O., Barton, W., Skuse, P., Penney, N. C., Garcia-Perez, I., Murphy, E. F., et al. (2018b). A Prospective Metagenomic and Metabolomic Analysis of the Impact of Exercise and/or Whey Protein Supplementation on the Gut Microbiome of Sedentary Adults. mSystems 3. doi: 10.1128/mSystems.00044-18.

De Almeida, M. L. M., Feringer, W. H., Jr., Carvalho, J. R. G., Rodrigues, I. M., Jordão, L. R., Fonseca, M. G., et al. (2016). Intense exercise and aerobic conditioning associated with chromium or L-carnitine supplementation modified the fecal microbiota of fillies. PLoS ONE 11. doi: 10.1371/journal.pone.0167108.

Dupuit, M., Rance, M., Morel, C., Bouillon, P., Boscaro, A., Martin, V., et al. (2022). Effect of Concurrent Training on Body Composition and Gut Microbiota in Postmenopausal Women with Overweight or Obesity. Med Sci Sports Exerc 54, 517–529. doi: 10.1249/MSS.0000000000002809.

Dziarski, R., Park, S. Y., Kashyap, D. R., Dowd, S. E., and Gupta, D. (2016). Pglyrp-Regulated Gut Microflora Prevotella falsenii, Parabacteroides distasonis and Bacteroides eggerthii Enhance and Alistipes finegoldii Attenuates Colitis in Mice. PLOS ONE 11, e0146162. doi: 10.1371/journal.pone.0146162.

Egan, B., and Zierath, J. R. (2013). Exercise metabolism and the molecular regulation of skeletal muscle adaptation. Cell Metab 17, 162–184. doi: 10.1016/j.cmet.2012.12.012.

Ghosh, S., Whitley, C. S., Haribabu, B., and Jala, V. R. (2021). Regulation of Intestinal Barrier Function by Microbial Metabolites. Cell Mol Gastroenterol Hepatol 11, 1463–1482. doi: 10.1016/j.jcmgh.2021.02.007.

Giacco, A., delli Paoli, G., Simiele, R., Caterino, M., Ruoppolo, M., Bloch, W., et al. (2020). Exercise with food withdrawal at thermoneutrality impacts fuel use, the microbiome, AMPK phosphorylation, muscle fibers, and thyroid hormone levels in rats. Physiol Rep 8, e14354. doi: 10.14814/phy2.14354.

Hoffman-Goetz, L., Pervaiz, N., and Guan, J. (2009). Voluntary exercise training in mice increases the expression of antioxidant enzymes and decreases the expression of TNF-alpha in intestinal lymphocytes. Brain Behav Immun 23, 498–506. doi: 10.1016/j.bbi.2009.01.015.

Hoffman-Goetz, L., Pervaiz, N., Packer, N., and Guan, J. (2010). Freewheel training decreases pro- and increases anti-inflammatory cytokine expression in mouse intestinal lymphocytes. Brain Behav Immun 24, 1105–1115. doi: 10.1016/j.bbi.2010.05.001.

Houghton, D., Stewart, C. J., Stamp, C., Nelson, A., Aj Ami, N. J., Petrosino, J. F., et al. (2018). Impact of Age-Related Mitochondrial Dysfunction and Exercise on Intestinal Microbiota Composition. Journals of Gerontology - Series A Biological Sciences and Medical Sciences 73, 571–578. doi: 10.1093/gerona/glx197.

Hughes, R. L., and Holscher, H. D. (2021). Fueling Gut Microbes: A Review of the Interaction between Diet, Exercise, and the Gut Microbiota in Athletes. Adv Nutr 12, 2190–2215. doi: 10.1093/advances/nmab077.

Ismail, A. S., Severson, K. M., Vaishnava, S., Behrendt, C. L., Yu, X., Benjamin, J. L., et al. (2011). Gammadelta intraepithelial lymphocytes are essential mediators of host-microbial homeostasis at the intestinal mucosal surface. Proc Natl Acad Sci U S A 108, 8743–8748. doi: 10.1073/pnas.1019574108.

Janabi, A. H. D., Biddle, A. S., Klein, D. J., and McKeever, K. H. (2017). The effects of acute strenuous exercise on the faecal microbiota in Standardbred racehorses. Comparative Exercise Physiology 13, 13–24. doi: 10.3920/CEP160030.

Janabis, A. H. D., Biddle, A. S., Klein, D., and McKeever, K. H. (2016). Exercise training-induced changes in the gut microbiota of Standardbred racehorses. Comparative Exercise Physiology 12, 119–130. doi: 10.3920/CEP160015.

Johnson, A. J., Zheng, J. J., Kang, J. W., Saboe, A., Knights, D., and Zivkovic, A. M. (2020a). A Guide to Diet-Microbiome Study Design. Frontiers in Nutrition 7. Available at: https://www.frontiersin.org/articles/10.3389/fnut.2020.00079 [Accessed June 20, 2023].

Johnson, E. L., Heaver, S. L., Waters, J. L., Kim, B. I., Bretin, A., Goodman, A. L., et al. (2020b). Sphingolipids produced by gut bacteria enter host metabolic pathways impacting ceramide levels. Nat Commun 11, 2471. doi: 10.1038/s41467-020-16274-w.

Kakiyama, G., Pandak, W. M., Gillevet, P. M., Hylemon, P. B., Heuman, D. M., Daita, K., et al. (2013). Modulation of the fecal bile acid profile by gut microbiota in cirrhosis. J Hepatol 58, 949–955. doi: 10.1016/j.jhep.2013.01.003.

Kern, T., Blond, M. B., Hansen, T. H., Rosenkilde, M., Quist, J. S., Gram, A. S., et al. (2020). Structured exercise alters the gut microbiota in humans with overweight and obesity—A randomized controlled trial. Int J Obes 44, 125–135. doi: 10.1038/s41366-019-0440-y.

Knight, R., Vrbanac, A., Taylor, B. C., Aksenov, A., Callewaert, C., Debelius, J., et al. (2018). Best practices for analysing microbiomes. Nat Rev Microbiol 16, 410–422. doi: 10.1038/s41579-018-0029-9.

Lamoureux, E. V., Grandy, S. A., and Langille, M. G. I. (2017). Moderate exercise has limited but distinguishable effects on the mouse microbiome. mSystems 2. doi: 10.1128/mSystems.00006-17.

Liu, X., Mao, B., Gu, J., Wu, J., Cui, S., Wang, G., et al. (n.d.). Blautia—a new functional genus with potential probiotic properties? Gut Microbes 13, 1875796. doi: 10.1080/19490976.2021.1875796.

Liu, Y., Wang, Y., Ni, Y., Cheung, C. K. Y., Lam, K. S. L., Wang, Y., et al. (2020). Gut Microbiome Fermentation Determines the Efficacy of Exercise for Diabetes Prevention. Cell Metabolism 31, 77–91.e5. doi: 10.1016/j.cmet.2019.11.001.

Lozupone, C. A., Stombaugh, J. I., Gordon, J. I., Jansson, J. K., and Knight, R. (2012). Diversity, stability and resilience of the human gut microbiota. Nature 489, 220–230. doi: 10.1038/nature11550.

Mahdieh, M., Maryam, J., Bita, B., Neda, F., Motahare, M., Mahboobeh, B., et al. (n.d.). A pilot study on the relationship between Lactobacillus, Bifidibactrium counts and inflammatory factors following exercise training. ARCHIVES OF PHYSIOLOGY AND BIOCHEMISTRY. doi: 10.1080/13813455.2021.1871763.

Mailing, L. J., Allen, J. M., Buford, T. W., Fields, C. J., and Woods, J. A. (2019). Exercise and the Gut Microbiome: A Review of the Evidence, Potential Mechanisms, and Implications for Human Health. Exercise and Sport Sciences Reviews 47, 75. doi: 10.1249/JES.0000000000000183.

Mattes, R. D., Rowe, S. B., Ohlhorst, S. D., Brown, A. W., Hoffman, D. J., Liska, D. J., et al. (2022). Valuing the Diversity of Research Methods to Advance Nutrition Science. Advances in Nutrition 13, 1324–1393. doi: 10.1093/advances/nmac043.

Meissner, M., Lombardo, E., Havinga, R., Tietge, U. J. F., Kuipers, F., and Groen, A. K. (2011). Voluntary wheel running increases bile acid as well as cholesterol excretion and decreases atherosclerosis in hypercholesterolemic mice. Atherosclerosis 218, 323–329. doi: 10.1016/j.atherosclerosis.2011.06.040.

Meng, Y., Chen, L., Lin, W., Wang, H., Xu, G., and Weng, X. (2020). Exercise Reverses the Alterations in Gut Microbiota Upon Cold Exposure and Promotes Cold-Induced Weight Loss. Front Physiol 11, 311. doi: 10.3389/fphys.2020.00311.

Mika, A., Treuren, W. V., González, A., Herrera, J. J., Knight, R., and Fleshner, M. (2015). Exercise Is More Effective at Altering Gut Microbial Composition and Producing Stable Changes in Lean Mass in Juvenile versus Adult Male F344 Rats. PLOS ONE 10, e0125889. doi: 10.1371/journal.pone.0125889.

Mitchell, C. M., Davy, B. M., Hulver, M. W., Neilson, A. P., Bennett, B. J., and Davy, K. P. (2019). Does Exercise Alter Gut Microbial Composition? A Systematic Review. Med Sci Sports Exerc 51, 160–167. doi: 10.1249/MSS.0000000000001760.

Moitinho-Silva, L., Wegener, M., May, S., Schrinner, F., Akhtar, A., Boysen, T., et al. (2021). Short-term physical exercise impacts on the human holobiont obtained by a randomised intervention study. BMC MICROBIOLOGY 21. doi: 10.1186/s12866-021-02214-1.

Mokhtarzade, M., Shamsi, M., Abolhasani, M., Bakhshi, B., Sahraian, M., Quinn, L., et al. (2021). Home-based exercise training influences gut bacterial levels in multiple sclerosis. COMPLEMENTARY THERAPIES IN CLINICAL PRACTICE 45. doi: 10.1016/j.ctcp.2021.101463.

Morita, E., Yokoyama, H., Imai, D., Takeda, R., Ota, A., Kawai, E., et al. (2019). Aerobic exercise training with brisk walking increases intestinal bacteroides in healthy elderly women. Nutrients 11. doi: 10.3390/nu11040868.

Motiani, K. K., Collado, M. C., Eskelinen, J.-J., Virtanen, K. A., Löyttyniemi, E., Salminen, S., et al. (2020). Exercise Training Modulates Gut Microbiota Profile and Improves Endotoxemia. Medicine and Science in Sports and Exercise 52, 94. doi: 10.1249/MSS.0000000000002112.

Oxford Centre for Evidence-Based Medicine: Levels of Evidence (March 2009) — Centre for Evidence-Based Medicine (CEBM), University of Oxford (n.d.). Available at: https://www.cebm.ox.ac.uk/resources/levels-of-evidence/oxford-centre-for-evidence-based-medicine-levels-of-evidence-march-2009 [Accessed January 20, 2023].

Packer, N., and Hoffman-Goetz, L. (2012). Exercise training reduces inflammatory mediators in the intestinal tract of healthy older adult mice. Can J Aging 31, 161–171. doi: 10.1017/S0714980812000104.

Parker, B. J., Wearsch, P. A., Veloo, A. C. M., and Rodriguez-Palacios, A. (2020). The Genus Alistipes: Gut Bacteria With Emerging Implications to Inflammation, Cancer, and Mental Health. Frontiers in Immunology 11. Available at: https://www.frontiersin.org/articles/10.3389/fimmu.2020.00906 [Accessed August 21, 2023].

Resende, A., Leite, G., and Junior, A. (2021). Changes in the Gut Bacteria Composition of Healthy Men with the Same Nutritional Profile Undergoing 10-Week Aerobic Exercise Training: A Randomized Controlled Trial. NUTRIENTS 13. doi: 10.3390/nu13082839.

Ribeiro, F. M., Ribeiro, C. F. A., Cláudia M. G. A., Castro, A. P., Almeida, J. A., Franco, O. L., et al. (2019). Limited effects of low-to-moderate aerobic exercise on the gut microbiota of mice subjected to a high-fat diet. Nutrients 11. doi: 10.3390/nu11010149.

Shanahan, E. R., McMaster, J. J., and Staudacher, H. M. (2021). Conducting research on diet-microbiome interactions: A review of current challenges, essential methodological principles, and recommendations for best practice in study design. J Hum Nutr Diet 34, 631–644. doi: 10.1111/jhn.12868.

Sun, S., Lei, O., Nie, J., Shi, Q., Xu, Y., and Kong, Z. (2022). Effects of Low-Carbohydrate Diet and Exercise Training on Gut Microbiota. FRONTIERS IN NUTRITION 9. doi: 10.3389/fnut.2022.884550.

Sun, S., Lulla, A., Sioda, M., Winglee, K., Wu, M. C., Jacobs, D. R., et al. (2019). Gut microbiota composition and blood pressure: The CARDIA study. Hypertension 73, 998– 1006. doi: 10.1161/HYPERTENSIONAHA.118.12109.

Taniguchi, H., Tanisawa, K., Sun, X., Kubo, T., Hoshino, Y., Hosokawa, M., et al. (2018). Effects of short-term endurance exercise on gut microbiota in elderly men. Physiological Reports 6. doi: 10.14814/phy2.13935.

Torquati, L., Gajanand, T., Cox, E. R., Willis, C. R. G., Zaugg, J., Keating, S. E., et al. (2022). Effects of exercise intensity on gut microbiome composition and function in people with type 2 diabetes. European Journal of Sport Science. doi: 10.1080/17461391.2022.2035436.

Tzemah Shahar, R., Koren, O., Matarasso, S., Shochat, T., Magzal, F., and Agmon, M. (2020). Attributes of Physical Activity and Gut Microbiome in Adults: A Systematic Review. Int J Sports Med 41, 801–814. doi: 10.1055/a-1157-9257.

Walshe, N., Cabrera-Rubio, R., Collins, R., Puggioni, A., Gath, V., Crispie, F., et al. (2021). A Multiomic Approach to Investigate the Effects of a Weight Loss Program on the Intestinal Health of Overweight Horses. FRONTIERS IN VETERINARY SCIENCE 8. doi: 10.3389/fvets.2021.668120.

Wang, K., Liao, M., Zhou, N., Bao, L., Ma, K., Zheng, Z., et al. (2019). Parabacteroides distasonis Alleviates Obesity and Metabolic Dysfunctions via Production of Succinate and Secondary Bile Acids. Cell Reports 26, 222–235.e5. doi: 10.1016/j.celrep.2018.12.028.

Wang, X., Hu, Y., Zhu, X., Cai, L., Farooq, M. Z., and Yan, X. (2023). Bacteroides-derived isovaleric acid enhances mucosal immunity by facilitating intestinal IgA response in broilers. J Anim Sci Biotechnol 14, 4. doi: 10.1186/s40104-022-00807-y.

Warbeck, C., Dowd, A., Kronlund, L., Parmar, C., Daun, J., Wytsma-Fisher, K., et al. (2021). Feasibility and effects on the gut microbiota of a 12-week high-intensity interval training plus lifestyle education intervention on inactive adults with celiac disease. APPLIED PHYSIOLOGY NUTRITION AND METABOLISM 46, 325–336. doi: 10.1139/apnm-2020-0459.

Wei, S., Brejnrod, A. D., Trivedi, U., Mortensen, M. S., Johansen, M. Y., Karstoft, K., et al. (2022). Impact of intensive lifestyle intervention on gut microbiota composition in type 2 diabetes: a post-hoc analysis of a randomized clinical trial. Gut Microbes 14. doi: 10.1080/19490976.2021.2005407.

Yang, H.-L., Li, M.-M., Zhou, M.-F., Xu, H.-S., Huan, F., Liu, N., et al. (2021a). Links Between Gut Dysbiosis and Neurotransmitter Disturbance in Chronic Restraint Stress-Induced Depressive Behaviours: the Role of Inflammation. Inflammation 44, 2448–2462. doi: 10.1007/s10753-021-01514-y.

Yang, W., Liu, Y., Yang, G., Meng, B., Yi, Z., Yang, G., et al. (2021b). Moderate-Intensity Physical Exercise Affects the Exercise Performance and Gut Microbiota of Mice. FRONTIERS IN CELLULAR AND INFECTION MICROBIOLOGY 11. doi: 10.3389/fcimb.2021.712381.

Zhong, X., Powell, C., Phillips, C., Millar, S., Carson, B., Dowd, K., et al. (2021). The Influence of Different Physical Activity Behaviours on the Gut Microbiota of Older Irish Adults. JOURNAL OF NUTRITION HEALTH & AGING 25, 854–861. doi: 10.1007/s12603-021-1630-6.

