## Supplemental Table 1 for "A Systematic Review on the Effects of Exercise on Gut Microbial Diversity, Taxonomic Composition, and Microbial Metabolites: Identifying Research Gaps and Future Directions"

**Supplemental Table 1**. Details of Search Methods

| **Database** | **Keywords** |
| --- | --- |
| PubMed | (“Exercise” [Mesh] AND (“Microbiota” [Mesh] OR "Gastrointestinal Microbiome"[Mesh]) |
| Web of Science | (Exercise OR Training OR “Physical Activity”) AND (Gut OR Gastro* OR Gastric OR Intestinal OR Fecal OR Cecal OR Faecal OR Enteric) AND (Microbiome OR Microbiota OR Microflora OR Microbiomes OR Microbe OR Microorganism OR Microbiology OR Microbial OR Flora OR Bacteria) |
| Cochrane Library | (Exercise OR Training OR "Physical activity") AND (Gut OR Gastro* OR Gastric OR Intestinal OR Fecal OR Cecal OR Faecal OR Enteric) AND (Microbiome OR Microbiota OR Microflora OR Microbiomes OR Microbe OR Microorganism OR Microbiology OR Microbial OR Flora OR Bacteria) |
| SCOPUS | TITLE-ABS-KEY ((Exercise OR Training OR "Physical activity") AND (Gut OR Gastro* OR Gastric OR Intestinal OR Fecal OR Cecal OR Faecal OR Enteric) AND (Microbiome OR Microbiota OR Microflora OR Microbiomes OR Microbe OR Microorganism OR Microbiology OR Microbial OR Flora OR Bacteria)) |

The databases (PubMed, Web of Science, Cochrane Library, and Scopus) were searched until October 29, 2021 and were then updated from October 2021 to June 1, 2022 by JD, JC, and SS. Articles were reviewed and screened by three authors (JD, JC, SS) based on established criteria.
